# DIVAID: Consistent division of atrial geometries from multimodal imaging according to the EHRA/EACVI 15-segment bi-atrial model

**DOI:** 10.64898/2026.04.22.26351448

**Authors:** Christian Goetz, Martin Eichenlaub, Kerstin Schmidt, Felix Wiedmann, Eric Invers Rubio, Patricia Martínez Díaz, Armin Luik, Till Althoff, Constanze Schmidt, Axel Loewe

## Abstract

The recently published EHRA/EACVI consensus statement on a standardized bi-atrial regionalization provides new opportunities for consistent regional analyses across patients, imaging modalities and clinical centers. To make this standardized regionalization widely accessible, we developed the open-source software DIVAID, which automatically divides bi-atrial geometries according to the proposed regions, ensuring consistency, reproducibility and operator independence. We evaluated the accuracy of the algorithm by comparing its results to manual expert annotations across 140 geometries from multiple modalities and centers. Veins were automatically clipped correctly in 81% and orifices annotated correctly in 100 % of cases. The median (interquartile range; IQR) Dice similarity coefficient (DSC) for left atrial regions was 0.98 (0.96 – 1.00) for DIVAID-expert and 0.98 (0.94 – 1.00) for inter-expert comparisons. For right atrial geometries, DSC was higher for DIVAID-expert than for inter-expert comparisons at 0.90 (0.80 – 0.95) and 0.88 (0.74 – 0.94), respectively. To assess the accuracy of regional boundaries, we computed the mean average surface distance (MASD) for boundaries derived from automatic or manual annotations. The median (IQR) MASD between DIVAID and experts was 0.17 mm (0.03 – 0.78) and 1.93 mm (0.65 – 3.96) in the left and right atrium, respectively. To conclude, DIVAID robustly divides anatomically diverse bi-atrial geometries according to the 15-segment model, while outperforming cardiac experts in both speed and consistency, and demonstrating an accuracy of regional boundaries comparable to the spatial resolution of cardiac imaging modalities. By providing automated, consistent atrial regionalization, DIVAID enables large-scale, standardized regional analyses and data-driven investigation of harmonized, multi-dimensional datasets, which may advance atrial arrhythmia research and personalized treatment strategies.

## 1. Introduction

High-resolution imaging and electroanatomical mapping (EAM) play a fundamental role in cardiac electrophysiology research and treatment planning for patients with arrhythmias. Recent technological advances open new opportunities for more precise and local analyses of cardiac morphology and function. However, standardized quantitative comparisons within and between subjects, as well as across imaging modalities require (i) a common reference system and (ii) an automated, operator- and modality-independent tool applying this reference system to the cardiac geometry. With respect to the left ventricle, a standardized regionalization, known as the 17-segment AHA model, was introduced more than two decades ago (Cerqueira et al., 2002), and has since been widely used in both clinical research (Abdel-Aty et al., 2004; Greenwood et al., 2012; Javadi et al., 2010; Ortiz-Pérez et al., 2008; Pereztol-Valdés et al., 2005) and computational modeling (Camps et al., 2024; Li et al., 2024). This was followed by the development of automated algorithms dividing the left ventricle in the proposed regions (Afshin et al., 2014; Bazhutina et al., 2023; Morris et al., 2023; Liang et al., 2015). Additionally, multiple software solutions aiming at a continuous parametrization of bi-ventricular geometries and their representation in two-dimensional (2D) space for easier interpretability and comparability were published. Paun et al. (2017) used solutions of Laplace’s equations to flatten left and right ventricular geometries to predefined reference frames. For the right ventricle, they used a half cylinder of unit height with a half disk as base, whereas for the left ventricle they used a cylinder of unit height with a unit disk as base for direct comparability with the 17-segment bull’s eye plot (Cerqueira et al., 2002). Moreover, to directly describe precise positions in any three-dimensional (3D) bi-ventricular geometry and to map data between bi-ventricular geometries, universal ventricular coordinates were introduced (Bayer et al., 2018). This approach was then further refined to enhance symmetry between the left and right ventricle and to ensure consistency among multiple geometries using normalized distances along bijective trajectories instead of solutions of Laplace’s equations (Schuler et al., 2021).

Several efforts have been made to adapt ventricular approaches to the atria. Similarly to Paun et al. (2017), surface flattening techniques were applied by Karim et al. (2014) who mapped the left atrium to a unit square, whereas Tobon-Gomez et al. (2015b) and Williams et al. (2017) mapped the left atrium (LA) to a disk. Since all these approaches required manual clipping of the veins and registering the 3D geometry to a 3D template before mapping to the 2D reference space, Nuñez-Garcia et al. (2020) enhanced this approach by semi-automatically clipping the veins and directly flattening the 3D geometry to a 2D disk, avoiding preprocedural registration. While 2D representations allow for easy and fast comparisons between multiple geometries and modalities, they inherently lose information about the 3D shape and introduce unavoidable distortions of the 3D geometry. To overcome this challenge, Roney et al. (2019) introduced universal atrial coordinates, in line with the proposition of the universal ventricular coordinates presented by Bayer et al. (2018). Using the solutions of Laplace’s equations, two coordinates per atrium are calculated, which facilitates image registration, as well as distortion-free 2D data visualization and interpretation. While universal atrial coordinates provide an important reference framework in the field, approaches based on solutions of Laplace’s equations may lead to inconsistencies across geometries and imaging modalities as noted by Schuler et al. (2021), particularly given the large anatomical variability of the atria. Notably, the methods for atrial parametrization presented above often rely on precise placement of manual seed points and/or manual preprocessing, such as clipping the veins and valves, which may limit reproducibility and operator independence.

Besides the efforts to standardize atrial parametrization by developing automated algorithms, many atrial regional studies have been performed (Assaf et al., 2023; Benito et al., 2018; Chen et al., 2013; Haissaguerre et al., 2014; Hunter et al., 2011; Nairn et al., 2020, 2023; Starreveld et al., 2018). However, such regionalizations previously described in literature were tailored to the specific modalities and analyses, lacking precise and reproducible definitions of regional boundaries. Lacking consensus of a common atrial reference system comparable to the 17-segment AHA model limits interpretability of the performed studies and applicability of the introduced algorithms.

The recently published clinical consensus statement on the EHRA/EACVI 15-segment bi-atrial model now opens new opportunities for standardized bi-atrial regionalizations (Althoff et al., 2025). It offers a standardized regionalization of both atria based on anatomical, electrophysiological and clinical considerations, and provides precise descriptions of regional boundaries, allowing for universal applicability and reproducibility. Moreover, the 15-segment biatrial model relies on the identification of anatomical landmarks in combination with standardized attitudinal axes and planes for precise and robust location of landmark points defining regional boundaries. However, given the complexity and large anatomical variability of atrial geometries with many varying sub-structures such as the pulmonary veins (PVs) (Prasanna et al., 2014) or the left atrial appendage (LAA) (García-Isla et al., 2018), also regarding different modalities, manual annotations can be time-consuming, operator- and modality-dependent and require advanced knowledge of cardiac anatomy and standardized orientations. To further standardize the regionalization of bi-atrial geometries across imaging modalities and to make the clinical consensus broadly accessible, we developed a framework that automatically, robustly, consistently and operator-, as well as modality-independently divides any biatrial geometry according to the proposed regions. Here, we introduce DIVAID (**DIV**ision of **A**tr**I**al **D**omains), the first software solution for standardized atrial bi-atrial regionalization aligned with the EHRA/EACVI 15-segment bi-atrial model. DIVAID extends approaches described above by providing a unified framework for 3D bi-atrial surface mesh preprocessing (e.g., automated vein clipping), as well as automated orifice annotation and landmark identification, enabling robust processing of geometries from multiple modalities including segmentations of tomographic images (Xiong et al., 2021) and EAMs (Bodagh and Williams, 2024). Released under an open-source license, our pipeline facilitates seamless application and integration into clinical studies and computational modeling.

Extending the proof-of-concept study by Goetz et al. (2024), we substantially enhanced our software across all three stages (Fig. 1) to improve robustness, usability, and clinical applicability. Preprocessing (stage 1) was completely revised to support both open and closed vein and valve configurations, and vein clipping was reworked to eliminate sensitivity to seed point placement, thereby increasing operator independence and robustness, with additional intersection checks for closely spaced veins. Automated orifice annotation (stage 2) was refined to further improve reliability. The division algorithm (stage 3) was redesigned to ensure full compliance with the EHRA/EACVI consensus statement and to enhance robustness and consistency across anatomically diverse geometries. Finally, validation was expanded by introducing evaluation of stages 1 and 2 and extending the division validation from 20 bi-atrial geometries from computed tomography (CT) and magnetic resonance imaging (MRI) assessed by a single expert to 140 anatomically diverse atrial geometries — including EAM — evaluated against three independent clinical experts, demonstrating improved robustness, consistency and generalizability.

**Figure 1:**
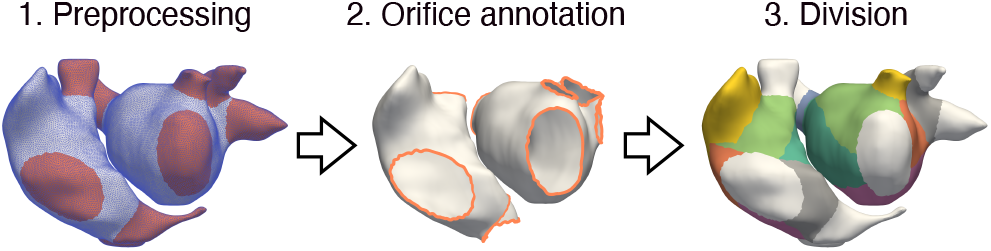
Overview of the individual stages of DIVAID.

## 2. Methods

To assure ease of use and broad compatibility, DIVAID is implemented as a Python-based, platform-independent framework under an open-source license, providing a versatile set of reusable functions for mesh and point-cloud manipulation (Goetz et al. (2026), https://gitlab.kit.edu/kit/ibt-public/divaid). Additionally, owing to its modular architecture, all scripts from stage 1 and stage 2 can be executed independently. As input, DIVAID accepts 3D mono- or bi-atrial surface meshes in VTK format with veins and valves optionally open or closed, derived from full resolution, full field of view data from volumetric imaging or EAM. Its output is a remeshed, watertight surface mesh, augmented with node and element data arrays containing the corresponding region tags. The individual stages of DIVAID are described in detail below.

### 2.1. Stage 1: Preprocessing

To minimize the impact of geometry- and modality-related variations on the regionalization procedure, the input geometry is preprocessed. This encompasses standardizing the surface mesh and clipping all veins and valves, as their orifices are important anatomical landmarks in the 15-segment bi-atrial model. These orifices are subsequently used to derive landmark points, which provide the foundation for defining the regional boundaries.

#### 2.1.1. Remeshing

First, all holes in the input mesh are closed using PyMeshLab to obtain a watertight mesh (Muntoni and Cignoni, 2021). This is essential to be able to compute reliable ray-mesh intersections, which will be used to clip the veins in the following. The surface mesh is resampled with a uniform edge length of 1 mm and smoothed using Humphrey’s Classes (HC) Laplacian smoothing to reduce noise and mitigate terracing artifacts caused by voxelation in clinical images of limited resolution, while preserving the original mesh volume (Vollmer et al., 1999), as shown in Fig. 2A. Scalar node and element data are mapped to the resampled mesh using a nearest neighbor algorithm. Nodes and elements in the resampled mesh with distances greater than 3 mm from any node or element in the original mesh are assigned NaN values to avoid introducing distortions.

**Figure 2:**
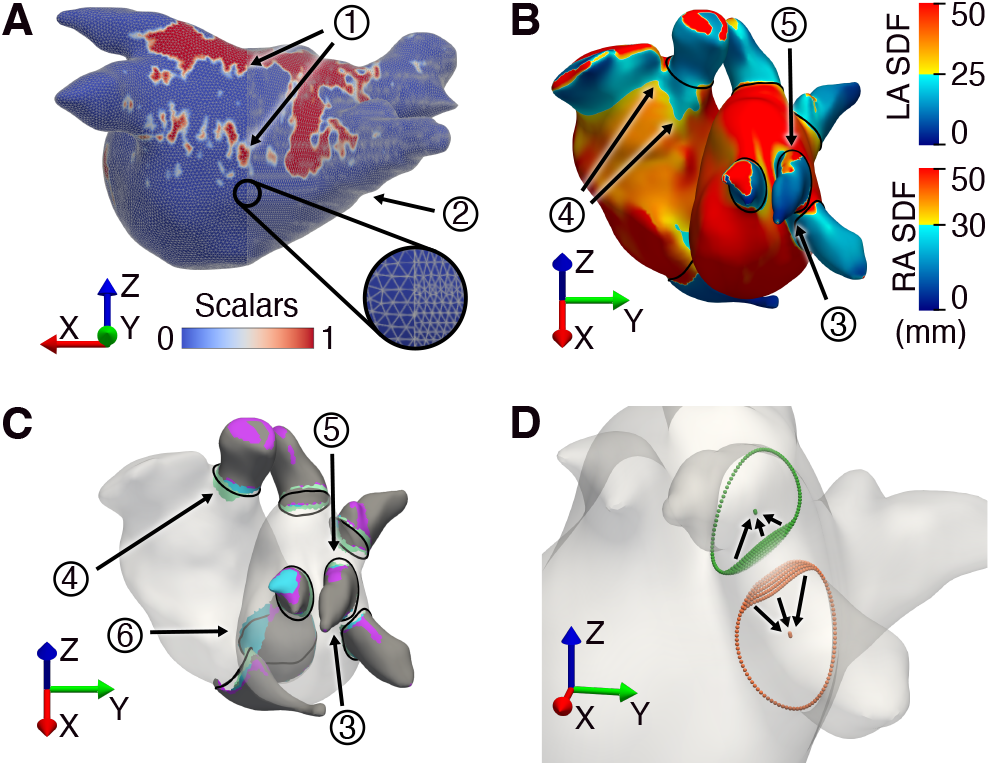
Individual steps of the preprocessing stage. (A) Left atrial input mesh (right) versus remeshed version (left). (1) Smooth transition of scalar values between the input and the remeshed mesh; (2) terracing artifacts. (B) Shape diameter function (SDF) values to clip the veins (blue) at the transition to the atrial body (yellow / red). Discrepancies from the anatomical orifices (black splines): (3) Distinct veins are connected to the same component; (4) proximal deviations due to thin non-tubular structures; (5) distal deviations due to imperfect tubular structures. (C) Separated veins after addressing (3) and (4) are shown in gray. Purple and blue elements denote additions from convex hull construction and planar clipping (case 5). Case (6) shows conversion of partially to fully encircling components after planar clipping. Elements added by region growing are shown in light green. (D) Intersection check between adjacent orifices. Spline points with distances below an iteratively increasing threshold are moved toward their respective spline centroids until two separate orifices are obtained.

#### 2.1.2. Valve clipping

The mitral valve (MV) and tricuspid valve (TV) are important anatomical landmarks of the atria. To extract their annuli, which serve as a basis for subsequent computation of regional boundaries, we distinguish between two scenarios. If the input mesh contains an orifice on which the distance between any two boundary nodes is larger than 30 mm, it is considered an annular ring. For each node on this boundary in the original mesh, the closest node in the resampled mesh is identified and a closed spline is fit to these nodes. Eventually, the spline is sampled at discrete points, and the nearest nodes in the resampled mesh are connected along the shortest geodesic paths. If the input mesh does not contain an orifice with the defined criteria, the annulus is defined manually. Seed nodes are interactively selected on the mesh to initiate the construction of a closed spline. The spline is updated with each newly selected seed node and transferred onto the mesh using the discrete nearest-node projection as described for the other scenario. The resulting clip can be previewed at any time prior to completion.

#### 2.1.3. Shape diameter function and seed placement

In addition to the MV and TV annuli, the 15-segment bi-atrial model relies on the vein ostia and the orifice of the LAA as characteristic landmarks. Specific points on these landmarks, as defined in the 15-segment bi-atrial model, are hereafter referred to as ‘landmark points’. Furthermore, the pulmonary veins (PVs), coronary sinus (CS), superior vena cava (SVC), inferior vena cava (IVC), and LAA are collectively referred to as ‘veins’ for simplicity. The openings of the PVs, CS, SVC and IVC are termed ‘ostia’, whereas those of the MV and TV are referred to as ‘annuli’. The opening of the LAA, as well as the collection of ostia and annuli is termed ‘orifice’. To robustly extract the transition from the veins to the atrial body, we use the shape diameter function (SDF) introduced by Shapira et al. (2008). Primarily designed to achieve consistent mesh partitioning, the SDF assigns a scalar value to each mesh node, representing the diameter of the object’s volume in its local neighborhood, as shown in Fig. 2B. In practice, for each node, the inward-pointing mean normal is computed from its neighboring elements and a ray-mesh intersection is calculated along this normal (ray casting). Since casting tens of thousands of rays directly in Python using the vectorized Möller and Trumbore (2005) algorithm is computationally expensive, we partitioned the rays into smaller batches and parallelized the computations. Substantial performance gains can be achieved by leveraging Intel’s high-performance ray tracing library, Embree (Wald et al., 2014). Even when executed on an ARM CPU via an emulator — since the Python wrapper for Embree is currently provided only for x86 architectures — ray-tracing operations ran more than 50 times faster than the native, vectorized, parallelized Python implementation. This improvement is primarily due to Embree’s optimized C++ implementation utilizing hardware acceleration. To ensure cross-platform compatibility and facilitate installation of DIVAID, Embree is employed whenever available, with the native Python implementation used as a fallback.

In parallel, seed nodes must be selected interactively on the veins that are to be clipped. This is required only to distinguish individual veins. As detailed below, the exact placement of the seeds does not aGect the resulting vein clipping.

#### 2.1.4. Vein clipping and clip verification

In the following, the SDF is employed to clip the veins at the transition to the atrial body and derive the corresponding orifices. First, the nodes with SDF values smaller than 25 mm in the LA and 30 mm in the right atrium (RA) are extracted, as shown in Fig. 2B. These values were motivated by multiple studies on the orifice diameter of the PVs, as well as of the vena cava and the LAA (Kato et al., 2003; Mačiulienė et al., 2018; Naksuk et al., 2016; Wittkampf et al., 2003). As illustrated in Fig. 2B, we observed three characteristic discrepancies between the splines representing the anatomically correct orifices (shown in black) and the boundary of the raw thresholded mesh (transition from blue to yellow): (3) Neighboring veins with closely spaced orifices remain connected to one component despite thresholding; (4) proximal deviation and connection to thin non-tubular structures of the atrial body; (5) distal deviation due to imperfect tubular shape of the vein. Distal deviations were primarily observed in the LA, as the PVs do not enter the atrial body perpendicularly, which leads to larger SDF values at vein nodes close to the atrial body. All three discrepancies will be specifically addressed in the following.

First, each seed node is assigned to the connected component of the thresholded mesh with the shortest Euclidean distance. If two seeds are assigned to the same connected component (3), distinct veins are separated using spectral clustering, applied on a symmetrized k-nearest neighbor graph. Next, potential connections to non-vein structures are removed. Since veins have a tubular shape, the respective element that intersects a node’s ray should be in the same connected component. If this is not the case, it might display a thin but non-tubular structure (4) and the respective nodes are removed from the component. The outcome of these two operations is highlighted in gray in Fig. 2C.

Next, to overcome discrepancies originating from distal deviations (5) and a closed spline is fit to the respective orifice to clip the veins from the atrial body. The first two steps focus on detaching element connections and removing outliers that are not considered as part of the respective vein, which may lead to holes in the vein meshes, especially towards the distal ends of the veins. To restore connectivity within respective veins, the convex hull of each vein is computed using its Delaunay triangulation and all enclosed nodes and elements are extracted. Elements added via the convex hull construction are highlighted in purple in Fig. 2C. For short veins, it may happen that the separation, applied to account for (3) and (4), leads to incompletely encircled connected components, as exemplified by (6) in Fig. 2C. Here, the gray component is limited to a half-cylindrical shape oriented toward the septal wall. Thus, a plane is fit to the normals of the boundary nodes of the incompletely encircled connected component using singular value decomposition. The normal of the plane is defined by the singular vector corresponding to the smallest singular value and the origin is set to the centroid of the boundary nodes. Elements on the distal side of this plane are added to the respective vein, as shown in blue in Fig. 2C.

After removing potential outliers, disconnecting non-tubular shapes from the veins and subsequently restoring connectivity within the veins, the current vein meshes represent the core of the veins with the entire distal parts. To determine the correct proximal location for clipping the veins, an iterative region-growing algorithm is applied, starting from the boundary nodes of each vein mesh. At each iteration, neighboring nodes of the current boundary nodes are evaluated. If at least 25 % of the neighboring nodes exhibit an SDF value below the defined threshold, the corresponding nodes are added to the mesh, as highlighted in light green in Fig. 2C. This prevents adding isolated outliers, while still allowing the incorporation of incomplete encircling parts that satisfy the inclusion condition. Thus, the final boundary is not necessarily parallel to the previously defined plane, enabling adaptation to veins that enter the atrium at non-orthogonal angles. The procedure terminates when the newly evaluated nodes no longer satisfy the inclusion criterion. Eventually, a smoothed, closed spline (shown in black) is fit to the boundary nodes located within 1.5 mm of the corresponding boundary of the raw, thresholded mesh.

Splines are projected to the mesh as described in Section 2.1.2 to obtain the orifices of the veins. To assure that adjacent veins with closely spaced orifices are not merged to one orifice, an intersection check is performed. In case an intersection is detected, the distance between the splines is increased iteratively. In each iteration, sampled points on both splines that are closer than an iteratively increasing distance threshold to points on the adjacent orifice spline are moved toward their respective spline centroids to satisfy the current imposed minimal distance constraint, as shown in Fig. 2D. This procedure is repeated until one separate orifice per seed node is obtained. For intersections between a vein orifice and a valve annulus, only the spline defining the vein orifice is adapted, as the valve annulus has already been defined in the previous stage. If the automatic procedure does not yield valid results or the geometry exhibits degenerate veins, the final vein clips can be adapted interactively, according to the previous stage by selecting seed nodes on the mesh that form a closed spline. The performance of the automatic vein clipping procedure is evaluated in Section 3.3.

### 2.2. Stage 2: Orifice annotation

In the following, all orifices are automatically annotated using agglomerative clustering and plane separation, inspired by Azzolin et al. (2023).

#### 2.2.1. Left atrium

In the LA, the orifice with the greatest number of nodes is assigned to the MV annulus, since it corresponds to the largest diameter. The remaining orifices are assigned to two clusters using proximity-based agglomerative clustering. The cluster that contains the orifice with the shortest Euclidean distance to the MV annulus is classified as the anatomical left (i.e., patient left), with the remaining cluster classified as the anatomical right. A plane defined by the mean of the centroids of each cluster and the centroid of the MV annulus separates superior from inferior orifices, as illustrated in Fig. 3A. The left superior orifice with the shortest Euclidean distance to the MV annulus is assigned to the LAA. This process is repeated with the remaining ostia and the superior ostia with the largest distances to the plane are assigned to the left superior PV (LSPV) and right superior PV (RSPV). Correspondingly, the inferior orifices with the largest distances to the plane are assigned to the left inferior PV (LIPV) and right inferior PV (RIPV), respectively. Besides the classic arrangement of four ostia, Ho et al. (2012) found specimen with five ostia, while others exhibited a common ostium on the left or right side. To account for this, any additional ostium between the most superior and most inferior vein ostium is assigned as middle ostium. In cases with a common trunk, the single ostium is defined as superior and inferior to allow for universal applicability of the definitions in the 15-segment bi-atrial model. Thus, the algorithm is able to process any LA geometry with one MV annulus, one LAA orifice and at least two ostia.

**Figure 3:**
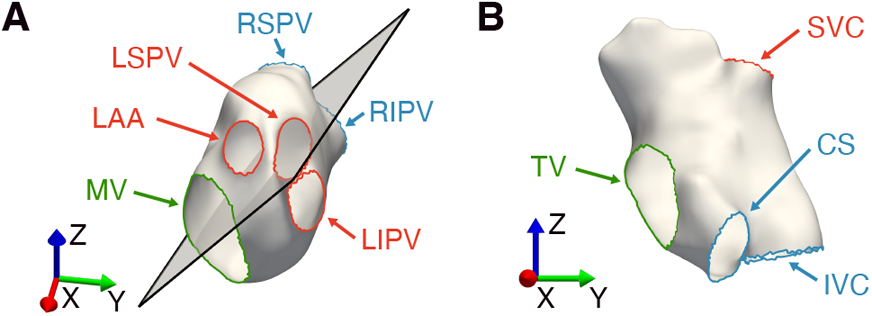
Automatic annotation of left and right atrial orifices. (A) The plane (black) defined by the centroid of the mitral valve (MV) annulus (green) and the centroids of the left (red) and right (blue) clusters separates superior and inferior orifices. LSPV/LIPV: left superior/left inferior pulmonary vein, RSPV/RIPV: right superior/right inferior pulmonary vein. (B) The tricuspid valve (TV) is shown in green, the cluster containing the superior vena cava (SVC) ostium in red, and the cluster containing the inferior vena cava (IVC) and the coronary sinus (CS) ostia in blue.

#### 2.2.2. Right atrium

Analogous to the LA, the RA orifice with the greatest number of nodes is assigned to the TV annulus. The remaining ostia are assigned to two clusters. Due to the proximity of the IVC and the CS ostia, the cluster that exhibits only one ostium corresponds to the SVC. Since the IVC ostium is significantly larger than the CS ostium, the ostium in the remaining cluster with the greatest number of nodes is assigned to the IVC (Klimek-Piotrowska et al., 2016). The remaining ostium corresponds to the CS, as shown in Fig. 3B.

### 2.3. Stage 3: Division

In the following, the identified anatomical landmarks will be used to divide atrial geometries according to the 15-segment bi-atrial model. Regional boundaries are defined by shortest geodesic paths between characteristic points on anatomical landmarks (landmark points), precisely defined using standardized body axes and planes.

#### 2.3.1. Body-centered anatomical coordinate system

To conform with established standards in medical imaging, we utilize a right-handed coordinate system with orthogonal unit vectors in which the x-axis increases towards the left (as opposed to the right) side of the patient, the y-axis increases towards the dorsal (as opposed to ventral) side of the patient and the z-axis increases towards the cranial (as opposed to caudal) end of the patient, as specified in DICOM PS3.3 Section C7.6.2.1.1 (National Electrical Manufacturers Association, 2025). This refers to a left-posterior-superior (LPS) coordinate system in contrast to the right-anterior-superior (RAS) coordinate system used in some medical software. Since atrial geometries from different imaging modalities may not always exhibit a body-centered anatomical LPS coordinate system, an approximation is inferred using anatomical landmarks.

For defining the x-axis of the LA, first, the vector from the mean centroid of the RPV ostia to the centroid of the LIPV ostium is computed. Since the RPV ostia are located more superiorly than the LPV ostia, this vector is rotated by –15° about the anteriorly oriented normal of the plane defined by the MV annulus centroid, the LIPV ostium centroid, and the mean centroid of the RPV ostia, using Rodrigues’ rotation formula (Rodrigues, 1840)), as shown in Fig. 4A. For the z-axis, the long cardiac axis is approximated by the vector from the centroid of the MV annulus to the arithmetic mean of the mean centroid of the LPV ostia and the mean centroid of the RPV ostia. This vector is then projected onto the plane orthogonal to the x-axis and subsequently rotated about the x-axis by 49.25° (Odille et al., 2017), as shown in Fig. 4B. The y-axis is obtained from the cross product of the z- and x-axes, completing a right-handed coordinate system. For defining the z-axis of the RA, we first compute the vector from the centroid of the IVC ostium to the centroid of the SVC ostium. To account for potential septal-lateral offsets between the ostia, this vector is projected onto the plane spanned by the normal of a plane fit to the TV annulus and the vector connecting the centroid of the TV annulus to the centroid of the SVC ostium (see Fig. 4C). For the y-axis, the vector connecting the centroid of the TV annulus to the centroid of the SVC ostium is first projected onto the plane orthogonal to the z-axis. As illustrated in Fig. 4D, this vector is then rotated by –60° about the z-axis. The x-axis is obtained from the cross product of the y- and z-axes, completing a right-handed coordinate system.

**Figure 4:**
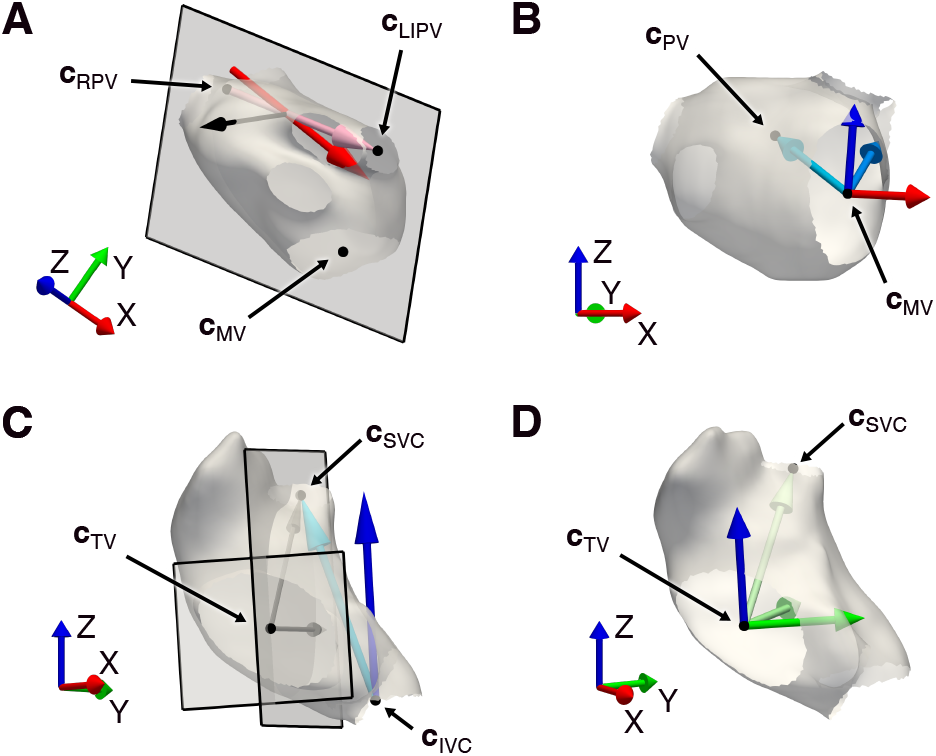
Definition of the body-centered anatomical coordi-nate system based on anatomical landmarks. (A) Left atrial x-axis (red): Vector between mean right pulmonary vein (RPV) and left inferior pulmonary vein (LIPV) centroids (light red), rotated by –15° about the anterior normal (black) of the plane through the mitral valve (MV), mean RPV, and LIPV centroids. (B) Left atrial z-axis (blue): Vector from the MV centroid to the arithmetic mean of the LPV and RPV centroids (light blue), projected onto the plane orthogonal to the x-axis (medium blue) and rotated by 49.25° about the x-axis. (C) Right atrial z-axis (blue): Vector between inferior vena cava (IVC) and superior vena cava (SVC) centroids (light blue), projected onto the plane defined by the tricuspid valve (TV) annulus normal (black) and the vector from the TV centroid to the SVC centroid (black). (D) Right atrial y-axis (green): Vector between TV and SVC centroids (light green), projected onto the plane orthogonal to the z-axis (medium green) and rotated by –60° about the z-axis. **c** denotes the centroid (arithmetic mean) of the nodes located on the ostium.

#### 2.3.2 Landmark points identification

Tab. 1 defines all landmark points underlying the 15-segment bi-atrial model, which constitute the basis for determining regional boundaries.

**Table 1.**
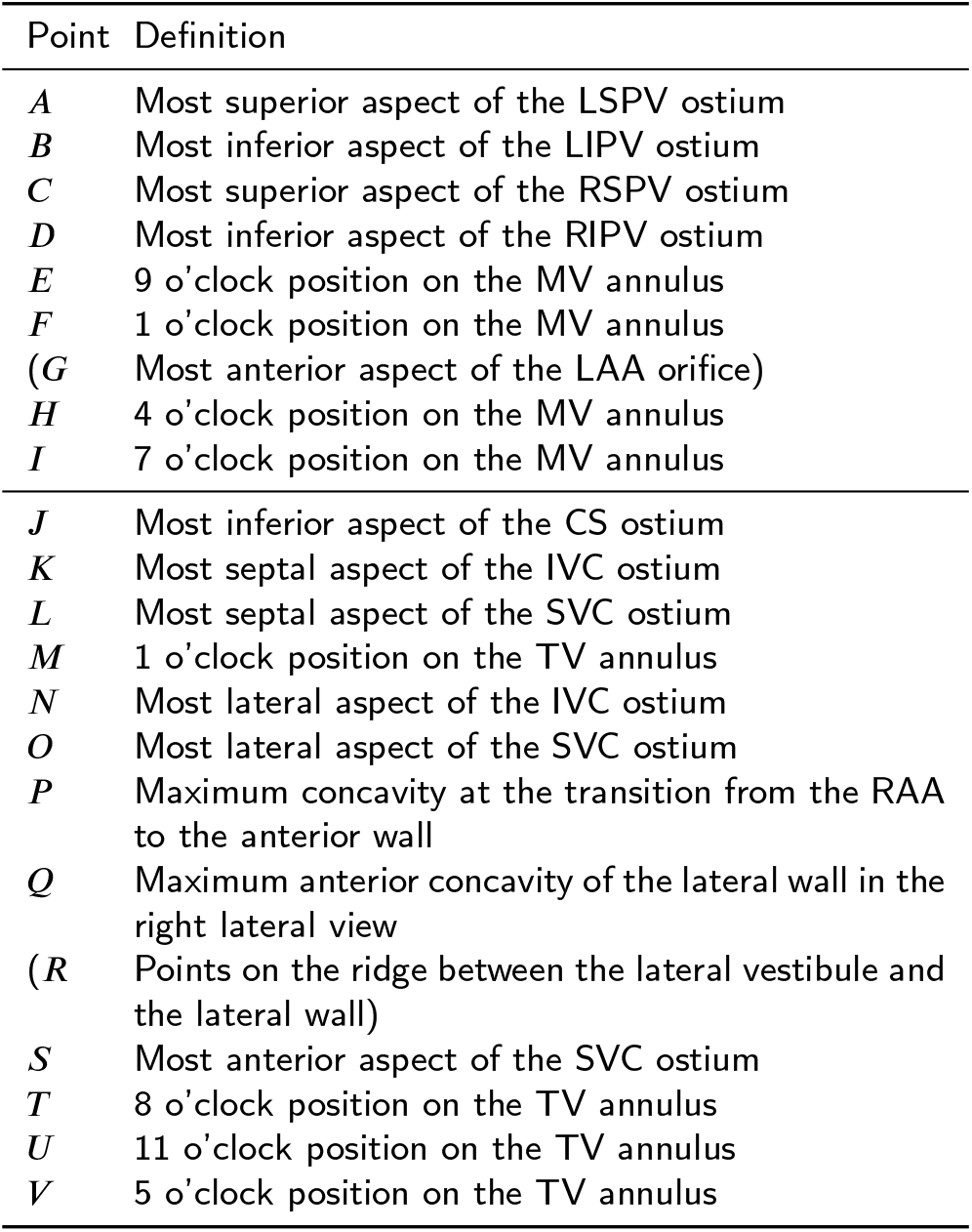
Location of all landmark points in the left (upper) and right (lower) atrium according to Althoff et al. (2025). Supporting points (in parentheses), not defined in the 15-segment bi-atrial model, yet required for regionalization. PV, pulmonary veins; L, left; R, right; S, superior; I, inferior; LAA, left atrial appendage; MV, mitral valve; CS, coronary sinus; IVC, inferior vena cava; SVC, superior vena cava; TV, tricuspid valve; RAA, right atrial appendage.

In the following, we detail how these landmark points are automatically identified. First, the preprocessed geometry is rotated into the body-centered anatomical coordinate system using rotation matrices constructed from the previously defined axes. All following operations are performed in this coordinate system. In the LA, landmarks *A* and *C* are defined as the nodes with the maximum z-coordinate, whereas landmarks *B* and *D* are defined by the minimum z-coordinate on the corresponding ostium. For landmarks *E, F, H* and *I*, the clock model introduced by Althoff et al. (2025) is used. The z-axis is projected onto the plane fit to the MV annulus, defining the 12 o’clock position, as shown in Fig. 5. Thus, landmarks *E, F, H* and *I* are obtained by rotating this vector about the apically directed normal vector of the MV plane by the respective angles (270° for 9 o’clock, 30° for 1 o’clock, 120° for 4 o’clock and 210° for 7 o’clock) and extracting the nodes with the smallest angular deviations. Additionally, for the lateral boundary of the anterior wall, the nodes on the LAA orifice with the shortest Euclidean distances to landmarks *A* and *F* are inferred. For subdivision of anatomical regions, the node on the LAA orifice with the shortest Euclidean distance to landmark *B* is determined. Landmark *G* is defined as the node on the LAA orifice with the minimum y-coordinate.

**Figure 5:**
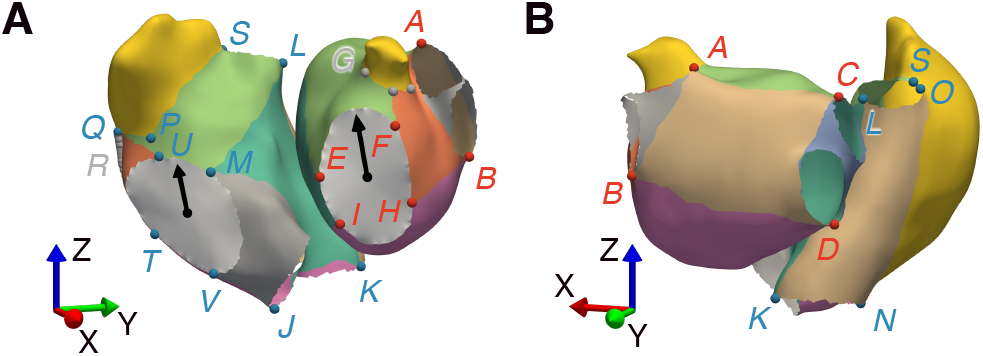
Characteristic landmark points, as defined in the 15-segment bi-atrial model, in the left (red) and right (blue) atrium, and supporting points (gray) not defined in the 15-segment bi-atrial model, yet required for regionalization. Veins are clipped, and valves are clipped and capped to improve visibility. Black arrows indicate the 12 o’clock position, originating from the respective valve centroids. (A) Left anterior oblique view (70°). (B) Posterior view rotated by –20° about the z-axis.

Analogous to the LA, landmarks *M, V, T* and *U* are determined by projecting the z-axis onto the plane fit to the TV annulus and rotating the projected vector about the apically directed normal vector of the plane by 30° for 1 o’clock, 150° for 4 o’clock, 240° for 8 o’clock, and 330° for 11 o’clock, respectively. Landmark *J* corresponds to the node on the CS ostium with the minimum z-coordinate. Landmark *S* corresponds to the node on the SVC ostium with the minimum y-coordinate. To obtain the most septal and lateral aspects of the SVC and IVC ostium, planes are fit to the respective ostia. Analogous to the clock model, the x-axis is then projected onto these planes and rotated about the superiorly directed normal vector by 45°and –45° to obtain the most septal (landmarks *L* and *M*) and lateral aspects (landmarks *N* and *O*), respectively.

Lastly, landmarks *P, Q* and *R* are computed. Since these landmark points define the boundary between the lateral vestibule and the lateral wall, we aim at extracting the prominent ridge at the junction between the lateral vestibule and the lateral wall, as explained by Althoff et al. (2025). First, candidate nodes are determined based on the following conditions. (i) The inferior and superior limits are defined by the z-coordinate of the 8 o’clock position on the TV (landmark *T* ) and the z-coordinate of the sum of the most superior aspect of the TV annulus and one sixth of the vector connecting the most superior aspect of the TV annulus to the centroid of the SVC ostium. (ii) The posterior limit is defined by a maximum distance to the TV plane of 20 mm. (iii) The septal limit is defined by a plane spanned by the normal vector of the plane fit to the TV annulus and the z-axis, with the origin set to the 8 o’clock position (landmark *T* ). Candidate nodes that satisfy these conditions are highlighted in blue in Fig. 6. Next, these nodes are separated into intervals of 2 mm along the z-axis. For each interval, 2D splines are fit to the xy-coordinates of nodes and placed at the interval midpoint along the z-axis, followed by principal component analysis of the sampled spline points. The first principal component corresponds to the long axis of the spline, approximately pointing from the point closest to the TV to the other end of the spline, as shown in the inset in Fig. 6A. Since most RA exhibit a prominent change in surface orientation from the lateral vestibule to the lateral wall, for each 2 mm interval, this ridge is approximated by extracting the sampled spline point with the maximum distal extent along the second principal component. Next, to account for potential outliers, a local coordinate system is defined, in which the x-axis is given by the mean of the vectors connecting the spline point closest to the TV to the previously identified high point, for each interval. The z-axis corresponds to the body-centered anatomical coordinate system and the y-axis is defined to complete a right-handed coordinate system. Quadratic polynomials were then fit in x-and y-directions, constraining the first polynomial to open towards the septal wall by enforcing a negative quadratic coemcient. The quadratic polynomials are evaluated at the midpoint of each z-axis interval satisfying (i) to generate point coordinates, as illustrated in Fig. 6B. Eventually, a smooth spline is fit, connecting the derived points on the ridge between the lateral vestibule and the lateral wall to the mid-path node of the shortest geodesic path between landmarks *T* and *N*. The spline is projected to the mesh, as described previously. Landmark *Q* corresponds to the most superior node of the projected spline, the remaining nodes refer to landmark *R*. Landmark *P* is identified as the node closest to the point located 15 mm septally from landmark *Q* along the normal vector of the plane spanned by the z-axis and the normal vector of the plane fit to the TV annulus. This approach provides a robust alternative to the definition used in the 15-segment bi-atrial model, which relies on identifying the point of maximum concavity at the transition from the right atrial appendage (RAA) to the anterior wall, a feature that may not be consistently detectable depending on the geometry and imaging modality.

**Figure 6:**
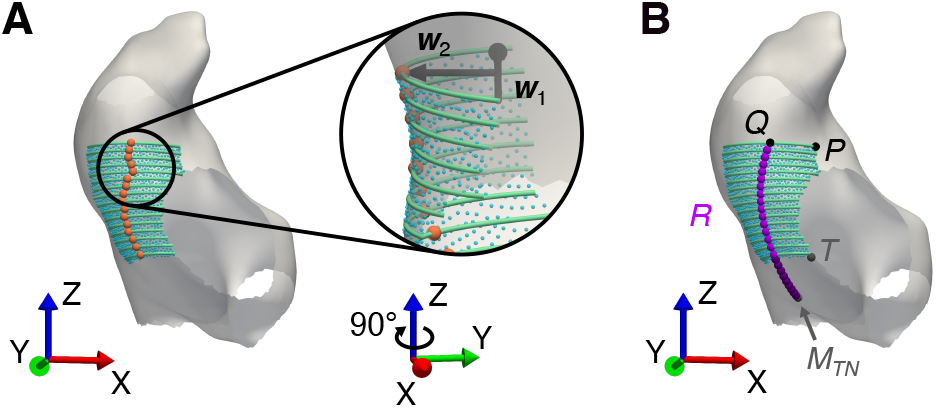
Identification of the prominent ridge between the lateral vestibule and the lateral wall in the right atrium. Constrained candidate nodes (blue) and splines fit to defined z-axis intervals (green). (A) Approximation of the ridge (orange) by extracting sampled spline points with maximum distal extent along the second principal component ***w***_2_. (B) Quadratic fits (purple) to the approximated ridge points (orange) assure a smooth boundary, with the lower segment (dark purple) extended to *M*_*TN*_ . Identification of landmark *P, Q* and *R* (black and purple), and supporting nodes (gray), where *M*_*TN*_ denotes the mid-node of the shortest geodesic path connecting landmarks *T* and *N*.

**Figure 7:**
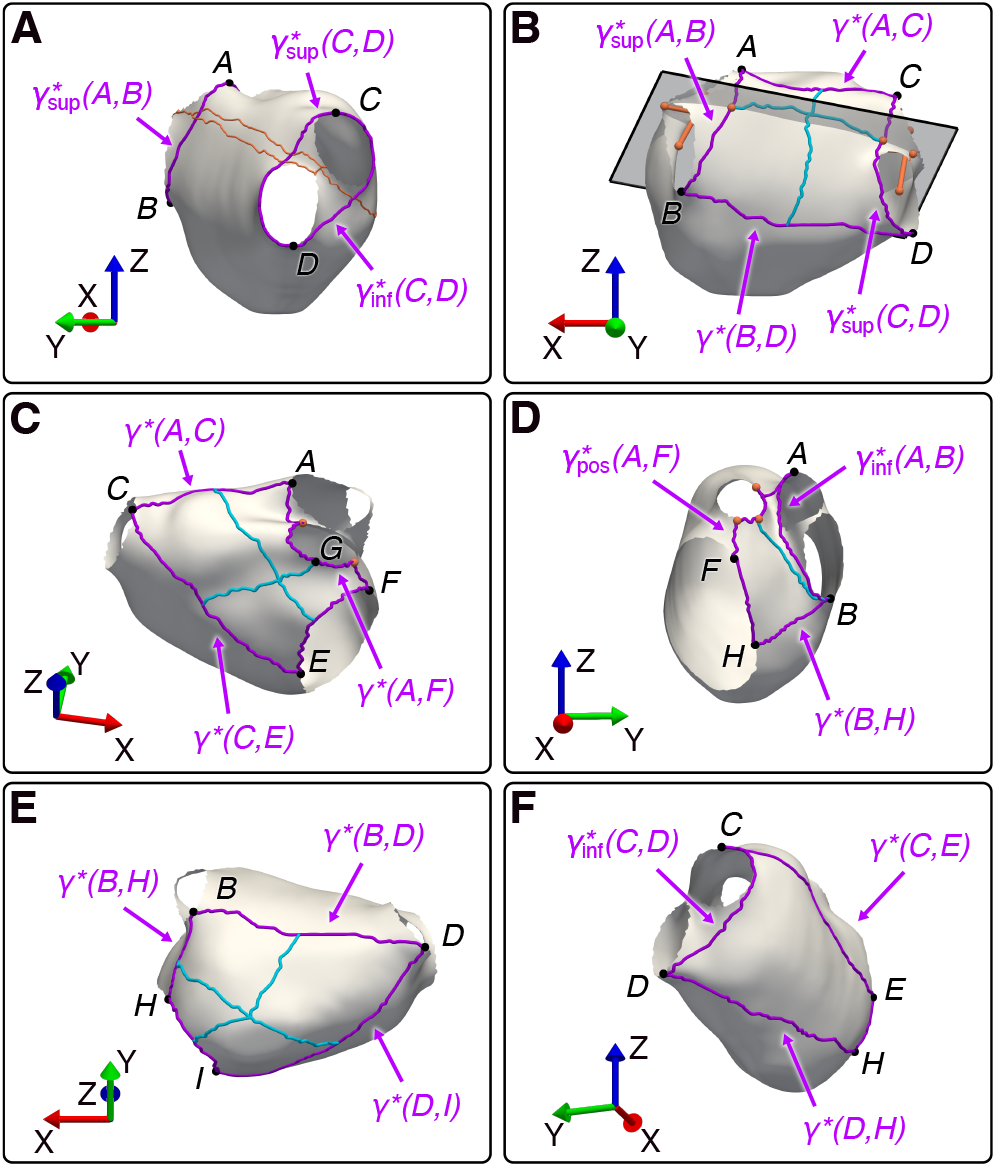
Region boundaries of the left atrium. (purple), subdividing boundaries (light blue) and supplementary points and paths (orange). (A) Left and right pulmonary venous antrum: Supplementary paths ensure that the shortest geodesic paths encircle the ostia superiorly and inferiorly. (B) Posterior wall: Orange nodes at the ostia indicate the corresponding centroids, while orange nodes between ostia represent mid-path nodes along the shortest geodesic path connecting neighboring ostia. Orange nodes on 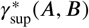 and 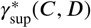 correspond to the nodes closest to the plane. (C) Anterior wall: Orange nodes indicate nodes on the left atrial appendage orifice closest (in Euclidean distance) to *A* and *F*, respectively. (D) Lateral wall: Orange nodes indicates nodes on the left atrial appendage orifice closest (in Euclidean distance) to *A* and *B*, respectively. (E) Inferior wall. (F) Septal wall.

#### 2.3.3. Boundaries computation

As detailed by Althoff et al. (2025), regional boundaries of the 15-segment bi-atrial model are determined by the respective shortest geodesic path between two landmark points. To obtain the shortest geodesic path *γ*^*^ (*Y, Z*) between two nodes *Y* and *Z* on an atrial surface mesh, we use Dijkstra’s algorithm (Dijkstra, 1959). In the following, we describe the boundaries of each region according to the numerical order of the segment numbers introduced by Althoff et al. (2025).

In the LA, the proximal boundaries of the pulmonary venous antrums are defined by the shortest circumference encircling the respective antrum and the ostia of the inferior and superior PVs on each side (AlthoG et al., 2025). Thus, the boundaries connecting landmarks *A* and *B*, and *C* and *D*, respectively, are separated into superior and inferior boundaries. For the superior boundaries, neighboring ostia on the same side are connected via the respective shortest geodesic paths. Additionally, each mid-path node is connected to the node on the MV annulus with the shortest Euclidean distance using the corresponding shortest geodesic path. Both are highlighted in orange in Fig. 7A. Temporarily removing these nodes along the respective paths from the mesh ensures that the superior boundaries 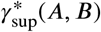 and 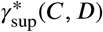 encircle the ostia superiorly. For the inferior boundaries, temporarily removing the nodes along the shortest geodesic paths connecting neighboring and opposing ostia ensures that the inferior boundaries 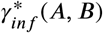and 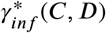encircle the ostia inferiorly. For the distal boundaries, the vein skeletons (i.e., the medial axes) are computed, using the SDF values following Shapira et al. (2008) and clip the veins orthogonally to the skeleton at a distance of 5 mm from the respective ostium.

To obtain the superior boundary *γ*^*^(*A, C*) and the inferior boundary *γ*^*^(*B, D*) of the posterior wall, landmarks *A* and *C* and landmarks *B* and *D* are connected using the corresponding shortest geodesic paths, respectively. The subdivision of the posterior wall into superior and inferior subregions aims at bisecting the carinas (AlthoG et al., 2025). Thus, we first compute the vectors between the centroids of neighboring ostia on the same side, as shown in Fig. 7B. The mean of these vectors is then projected onto the direction orthogonal to the vector connecting the mid-path nodes of the neighboring-ostium paths, yielding the normal of the fit plane. Then, the nodes of the boundaries 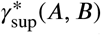 and 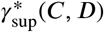 that are closest to this plane are connected along the shortest geodesic path, as shown in light blue in Fig. 7B. To subdivide the posterior wall into lateral and septal subregions, we compute the shortest geodesic paths bisecting *γ*^*^(*A, C*), the superior-inferior subdividing path, and *γ*^*^(*B, D*). The subregion labels a, b, c and d are defined according to a superior-inferior and lateral-septal orientation across all regions (AlthoG et al., 2025). The same convention is used for naming the regional boundaries.

The septal boundary of the anterior wall is defined by the shortest geodesic path *γ*^*^(*C,E*) connecting landmarks *C* and *E*. The MV annulus determines the inferior boundary. For the superior section of the lateral boundary, landmark *A* is connected to the node on the LAA orifice nearest to *A* in Euclidean distance via the shortest geodesic path. Similarly, for the inferior section, landmark *F* is connected to the node on the LAA orifice nearest to *F* in Euclidean distance via the shortest geodesic path. Both sections are joined with the anterior aspects of the LAA orifice to form *γ*^*^(*A, F* ), as shown in Fig. 7C. To subdivide the anterior wall into superior and inferior subregions, landmark *G* is connected to the mid-path node of *γ*^*^(*C, E*) via the shortest geodesic path. The lateral and septal subregions are obtained by computing the shortest geodesic paths bisecting *γ*^*^(*A, C*), the superior-inferior subdiving path, and *γ*^*^(*E, F* ).

The boundary of the LAA is defined by its transition to the atrial body, i.e. by its orifice, as defined in Section 2.1.4. The shortest geodesic path *γ*^*^(*B, H*) between landmarks *B* and *H* defines the infero-posterior boundary of the lateral wall. The apical boundary is determined by the MV annulus. To separate the left lateral ridge (segment 6a) from the lateral wall (segment 6b), the shortest geodesic path between landmark *B* and the node on the LAA orifice nearest to *B* in Euclidean distance is computed (see Fig. 7D).

The septal boundary of the inferior wall is defined by the shortest geodesic path *γ*^*^(*D, I*) connecting landmarks *D* and *I*. Inferiorly, the inferior wall is determined by the MV annulus. Similarly to the posterior and anterior wall, the subdivision of the inferior wall into superior and inferior subregions is defined by the shortest geodesic path bisecting *γ*^*^(*B, H*) and *γ*^*^(*D, I*). To subdivide the inferior wall into lateral and septal subregions, we compute the shortest geodesic paths bisecting *γ*^*^(*B, D*), the superior-inferior subdividing path, and *γ*^*^(*H, I*), as illustrated in Fig. 7E. All boundaries of the septal wall have already been introduced and are shown in Fig. 7F.

Regarding the RA, boundaries *γ*^*^(*K, J* ), *γ*^*^(*L, K*) and *γ*^*^(*L, M*) of the septal wall are defined by the respective shortest geodesic paths. To ensure, *γ*^*^(*M, J* ) encircles the CS ostium posteriorly, the nodes along the shortest geodesic path connecting the CS ostium to the TV annulus are temporarily removed from the mesh before the shortest geodesic path between landmarks *M* and *J* is computed (see Fig. 8A). The lateral boundary *γ*^*^(*O, N*) of the posterior venous wall is defined by the shortest geodesic path connecting landmarks *O* and *N*. As shown in Fig. 8B, the inferior and superior boundaries are determined by the ostia of the IVC and SVC, respectively.

**Figure 8:**
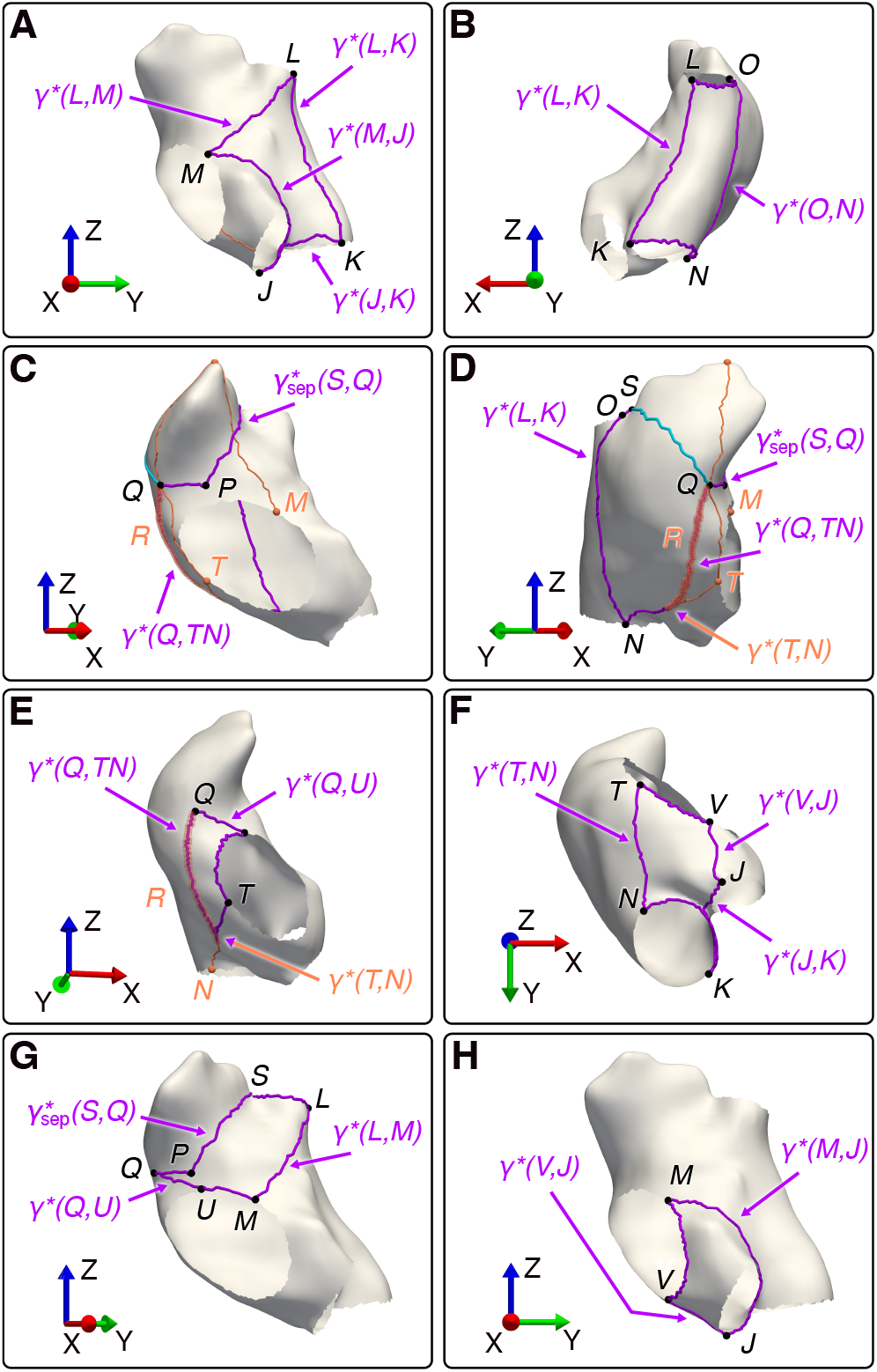
Region boundaries of the right atrium. (purple), subdividing boundaries (light blue) and supplementary points and paths (orange). (A) Septal wall: Supplementary path ensures that *γ*^*^(*M, J* ) encircles the coronary sinus ostium posteriorly. (B) Posterior wall. (C-D) Right atrial appendage (RAA) and lateral wall: Supplementary path connecting the most superior aspect of the RAA ridge to *M* ensures that 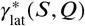 (light blue) encircles the RAA laterally. Supplementary path connecting the most superior aspect of the RAA ridge to *T* via *Q* ensures that 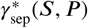 encircles the RAA septally. *Q* is connected to the mid-path node of *γ*^*^(*T, N*), passing through *R*. (E) Lateral vestibule. (F) Cavotricuspid isthmus. (G) Anterior wall. (H) Koch’s triangle.

The shortest geodesic path *γ*^*^(*S, P* ) defines the anterior boundary of the RAA and the lateral wall by connecting landmarks *S* and *P* . To ensure this path encircles the RAA septally, the nodes along the shortest geodesic path connecting the most superior aspect of the RAA ridge to landmark *T*, via landmark *Q*, are temporarily removed from the mesh. The path *γ*^*^(*S, P* ) is joined with the shortest geodesic path *γ*^*^(*P, Q*) connecting landmarks *P* and *Q* to form 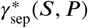, as shown in Fig. 8C, since landmarks *P* and *Q* are inter-dependent. The inferior boundary of the RAA and the lateral wall is determined by the shortest geodesic path *γ*^*^(*T, N*) connecting landmarks *T* and *N*. Towards the lateral vestibule, landmark *Q* is connected to the mid-path node of *γ*^*^(*T, N*) via the shortest geodesic paths *γ*^*^(*Q, T N*) passing through landmarks *R*, as shown in Fig. 8D. To separate the RAA (segment 11a) from the lateral wall (segment 11b), landmark *S* is connected to landmark *Q* laterally via the shortest geodesic path. To ensure this path encircles the RAA laterally, the nodes along the shortest geodesic path connecting the most superior aspect of the RAA ridge to landmark *M* are temporarily removed from the mesh.

The superior boundary of the lateral vestibule is defined by the shortest geodesic path *γ*^*^(*Q, U* ) connecting landmarks *Q* and *U*, as illustrated in Fig. 8E.

The septal boundary of the cavotricuspid isthmus is determined by the shortest geodesic path *γ*^*^(*V, J* ) between landmarks *V* and *J* (see Fig. 8F). The boundaries of the anterior wall and Koch’s triangle have already been introduced and are shown in Fig. 8G-H.

#### 2.3.4 Region assignment

Based on these region boundaries, region tags are assigned to all nodes and elements, following the segment numbering defined by Althoff et al. (2025). Node region tags are obtained by removing the corresponding boundary nodes from the mesh, extracting the largest connected component and computing the difference between the nodes of this component and those of the complete watertight mesh. Thus, boundary nodes separating adjacent regions may initially be assigned to multiple regions. This indirect assignment strategy is employed to preserve element connectivity within each region, particularly in thin anatomical sections or where boundaries merge at acute angles, as shown in Fig. 9. Element region tags are assigned based on the region tag shared by all nodes defining the element. Eventually, for regional boundaries separating two adjacent regions, the associated boundary nodes are assigned to exactly one of the two regions, ensuring that each node carries a single region tag. For further details on anatomical precedence, refer to the code (Goetz et al., 2026).

**Figure 9:**
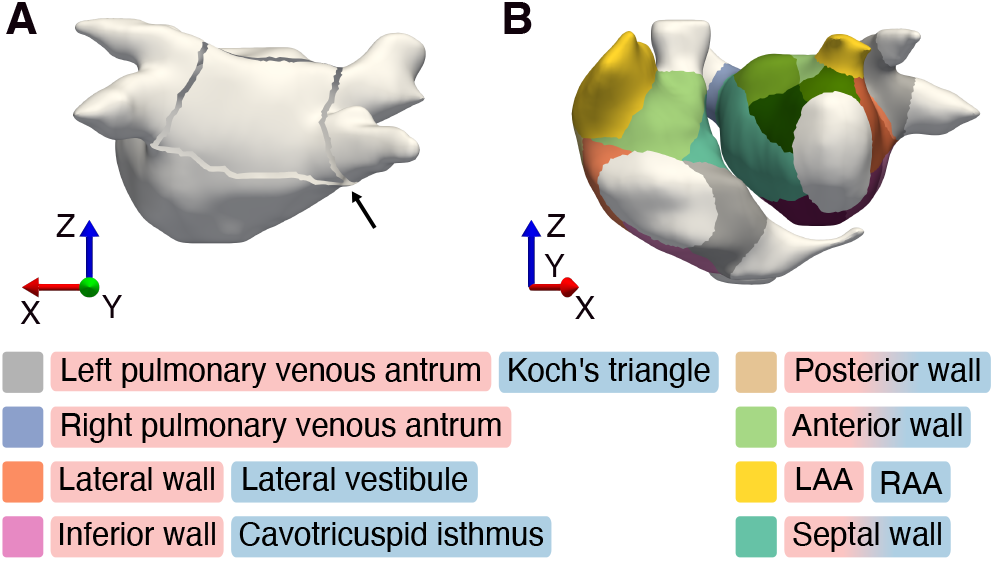
Result of the automated regionalization. (A) Exemplary region assignment of the left atrial posterior wall. Regional boundary nodes are removed from the mesh, the largest connected component is extracted, and the difference to the complete watertight mesh is computed. The black arrow highlights a case where acute boundary merging would incorrectly prevent nodes from being assigned to the region when using a direct assignment approach. (B) Final division applied to both atria including subdivisions.

### 2.4 Dataset

To evaluate DIVAID’s practical applicability and generalizability, we analyzed 140 atrial geometries collected from two centers, encompassing three imaging modalities: Center 1 provided 35 bi-atrial MRIs and 5 bi-atrial CTs, while center 2 contributed 20 bi-atrial EAMs and 20 LA MRIs. Both datasets exhibited substantial anatomical variability, including LAs with three, four or five PVs. The study was conducted in accordance with the Declaration of Helsinki, was approved by the institutional ethics committees and all patients provided written informed consent for research purposes prior to enrollment. MRIs were acquired on 3T scanners with a voxel size of 1.25 mm x 1.25 mm x 2.5 mm (reconstructed to 0.625 mm x 0.625 mm x 1.25 mm). Atrial segmentations were performed by two independent expert core laboratories: Adas group (Adas3D Medical SL, Barcelona, Spain) for MRIs from center 1 and Merisight (Marrek Inc., Sandy, UT, USA) for MRIs from center 2. Cardiac CT images were acquired using a SOMATOM Force scanner (Siemens Healthineers, Erlangen, Germany) at 90 kV with a field of view of 532 mm x 343 mm and pixel magnitude of 1:2. EAMs were obtained using the CARTO-3 mapping system (Biosense Webster, Diamond Bar, CA, USA) and a 20-polar Lasso-Nav mapping catheter or a PentaRay catheter (Nairn et al., 2023). Enrolled patients had a history of atrial fibrillation, which may lead to atrial structural remodeling, particularly enlargement of the left atrium (Nattel and Harada, 2014).

### 2.5. Validation

The performance of DIVAID was evaluated across all stages. Computation time was assessed using the Rosetta emulator on a MacBook Pro with an M2 Pro CPU (10 cores) to leverage Intel’s high-performance ray-tracing library. The annotation of the orifices was evaluated by an expert according to established anatomical standards and nomenclature. The performance of the vein clipping procedure was assessed visually by the same expert. Vein clips that did not accurately represent the transition between vein and atrial body were adapted manually using DIVAID’s interactive tool. Valves were clipped prior to pipeline execution using DIVAID’s interactive tool. Eventually, to validate the accuracy of the regionalization produced by DIVAID, we compared the regions and boundaries identified automatically with manual annotations from three independent experts (two electrophysiologists and one computational cardiac scientist). The experts interactively selected the landmark points defined by Althoff et al. (2025) on the clipped mesh, i.e., the output from stage 1 of DIVAID. Regional boundaries were then obtained automatically according to Section 2.3.3. For each atrium, a random subset was selected from the full dataset: five MRIs and five CTs from center 1, and five EAMs plus — for the LA — five additional MRIs from center 2, as shown in Fig. 10. These geometries were annotated by all experts to investigate inter-rater variability. In addition, for each atrium, ten MRIs from center 1 and five MRIs plus — for the LA — five additional MRIs from center 2 were randomly assigned to each expert. Thus, each expert annotated the multi-annotator subset consisting of 35 geometries (20 LA and 15 RA), as well as an additional 20 LA and 15 RA geometries of the single-annotator subset. Validation metrics were selected using the Metrics Reloaded toolkit (Maier-Hein et al., 2024) (see Section 6.3 for details): Dice similarity coemcient (DSC) as overlap-based metric (Eq. 1) and mean average surface distance (MASD) as boundary-based metric (Eq. 3). The DSC is defined as

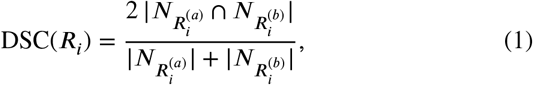

where *R*_*i*_ denotes the respective region, and 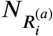 and 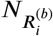 are the sets of nodes assigned to region *R*_*i*_ by annotators *a* and *b*, respectively. The annotators *a* and *b* may represent either automatic or manual annotations. The MASD is computed as follows:

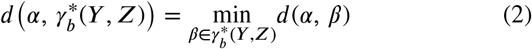

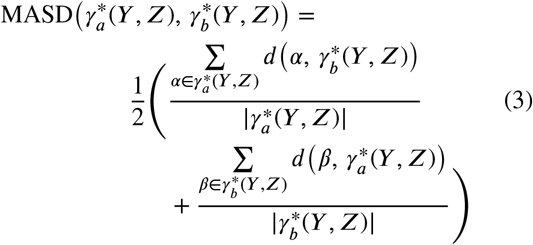

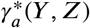 and 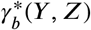denote the sets of nodes along the shortest geodesic paths connecting nodes *Y* and *Z* as placed by annotators *a* and *b*, respectively. For each node 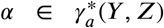, we compute the shortest Euclidean distance to any node in 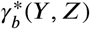. This process is repeated for each node in 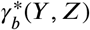. Taking the average of the means of these minimal distances in both directions yields the MASD between the boundaries derived by annotators *a* and *b*. Values are reported as median (interquartile range, IQR; 25th–75th percentile) unless stated otherwise.

**Figure 10:**
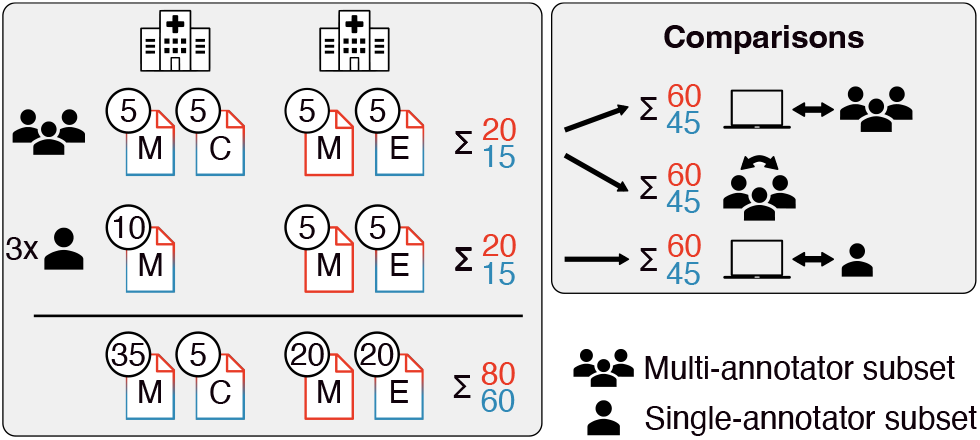
Validation dataset and comparisons. Left panel: Distribution of geometries into a multi-annotator subset (each geometry independently annotated by three experts; first row) and a single-annotator subset (each geometry annotated by one expert; second row). Columns correspond to geometries acquired at center 1 and center 2, respectively. Left atrial geometries are shown in red and right atrial geometries in blue. Right panel: Comparisons between automatic and manual annotations for both subsets, and inter-annotator comparisons within the multi-annotator subset. M, magnetic resonance imaging; C, computed tomography; E, electroanatomical mapping.

## 3. Results

### 3.1. Anatomical variability

The processed geometries showed substantial anatomical variability, with surface areas ranging from 72 cm^2^ to 214 cm^2^ in the LA and from 80 cm^2^ to 241 cm^2^ in the RA, and chamber volumes ranging from 42 mL to 245 mL in the LA and from 43 mL to 247 mL in the RA. As illustrated in Fig. 11, median (IQR) surface areas were comparable between the LA and RA, measuring 142 cm^2^ (118 – 162) and 143 cm^2^ (120 – 175), respectively. LA volumes were slightly higher than RA volumes: 125 mL (91 – 149) vs. 113 mL (87 – 155). All measurements were performed on atrial bodies with clipped veins and LAAs, with the resulting openings planarly capped. Among the 80 processed LA geometries, 74 geometries exhibited 4 PV ostia, 5 exhibited 3 PV ostia and one exhibited 5 PV ostia.

**Figure 11:**
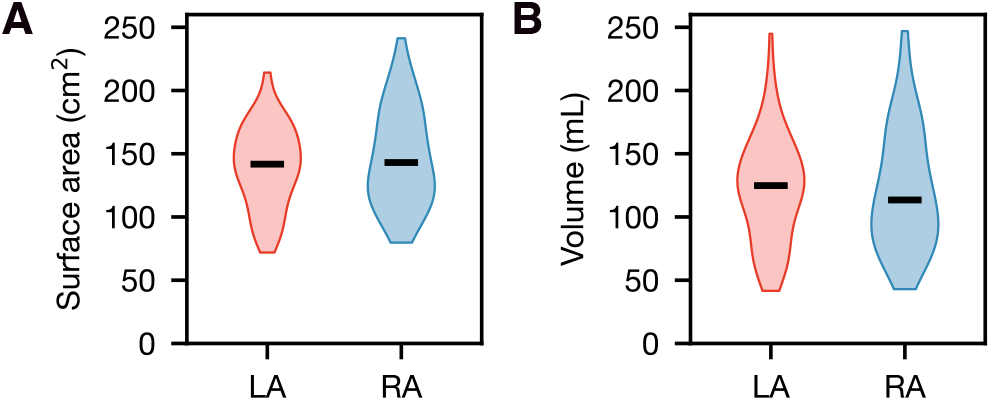
Surface area and chamber volume of the processed geometries. LA, left atrium; RA, right atrium.

### 3.2 Computation time

To evaluate DIVAID’s applicability for large-scale atrial regionalization, we analyzed its computation time and compared it with manual annotation. Fig. 12 summarizes the computation time for each stage of DIVAID. The computation time for the entire pipeline was 23.37 s (19.71 – 28.43) per geometry. Stage 1 required 21.48 s (18.43 – 25.42), stage 2 0.13 s (0.12 – 0.15) and stage 3 1.87 s (0.79 – 2.76). Seed placement and vein clipping were the largest contributors to stage 1 and showed higher runtimes in the LA than in the RA: 10.45 s (9.02 – 12.16) vs. 9.47 s (7.92 – 11.54) and 6.69 s (5.47 – 9.46) vs. 3.16 s (1.92 – 6.44). Similarly, stage 3 runtimes were higher for LA geometries than for RA geometries: 2.62 s (2.06 – 3.62) vs. 0.72 s (0.60 – 0.93). SDF calculation and orifice annotation were fastest, with computation times below 0.20 s. Compared to manual annotation, automated identification of landmark points was substantially faster, with runtimes of 0.01 s (0.005 – 0.07) vs. 131.42 s (104.11 – 186.54) per geometry.

**Figure 12:**
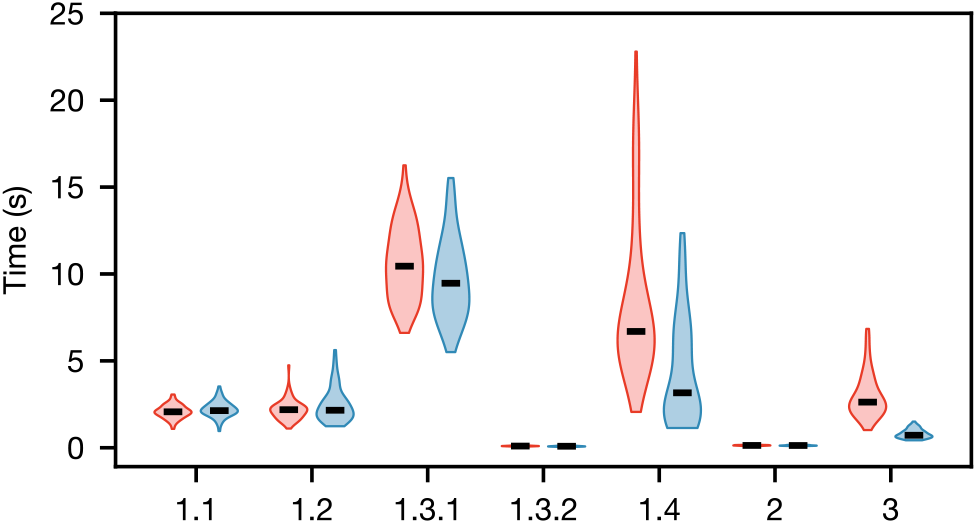
Computation time for the individual stages of DIVAID, shown separately for left atria (red) and right atria (blue). 1.1, remeshing; 1.2, valve clipping; 1.3.1, seed placement; 1.3.2, shape diameter function computation; 1.4, vein clipping; 2, orifice annotation; 3, division.

### 3.3 Vein clipping

The performance of the automated vein clipping procedure is illustrated in Fig. 13. In total, 452 of 558 visible veins (81 %) were clipped at the transition from the vein to the atrial body. Here, *visible* refers to veins that protrude from the atrial body and are therefore identifiable for automated clipping. In 15 cases, RA geometries exhibited no CS. Thus, these ostia were extracted manually based on their probable location, as shown in Supplementary Fig. 18A. The same procedure was performed for three LA geometries which exhibited no LSPV or RIPV (see Supplementary Fig. 18B). Note that these cases do not correspond to geometries with a common trunk; instead, the respective veins were not captured by the imaging modality.

**Figure 13:**
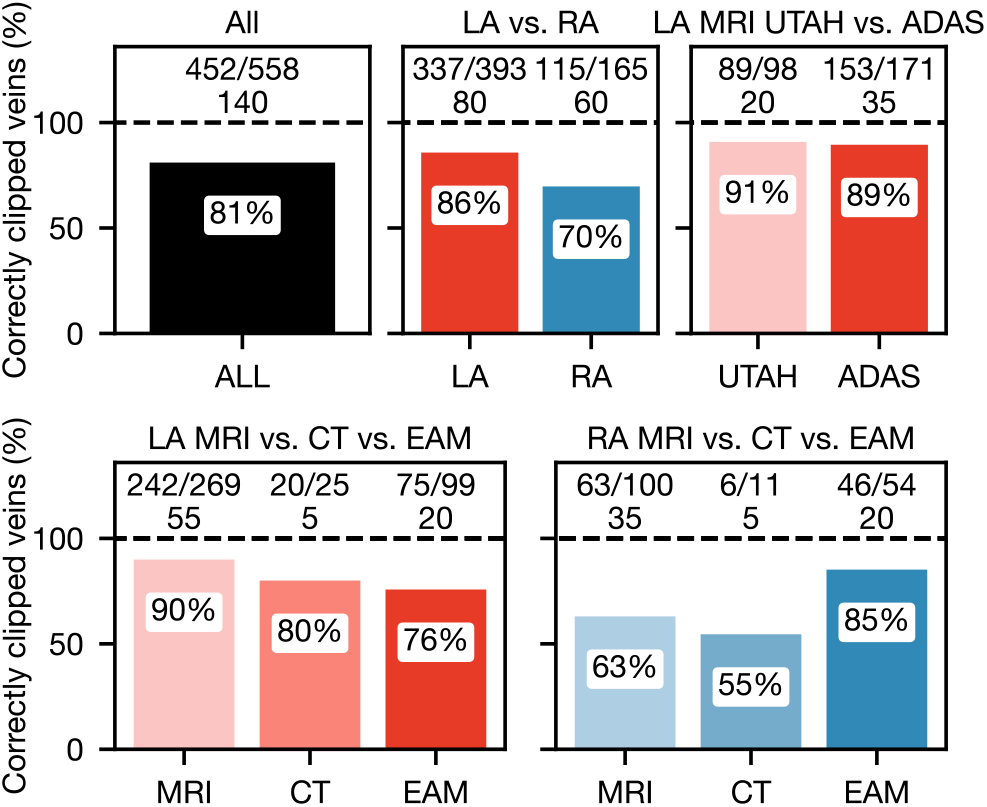
Validation of the automated vein clipping pro-cedure. In each panel, the upper row corresponds to the number of correctly clipped veins and the lower row to the number of geometries. LA, left atrium; RA, right atrium; MRI, magnetic resonance imaging; CT, computed tomography; EAM, electroanatomical mapping.

The automated vein clipping procedure performed better in the LA than in the RA (86 % vs. 70 %). While we observed a decline in performance from MRI to EAM in the LA (90 % vs. 76 %), performance was highest for EAM in the RA (85 %). We did not observe substantial discrepancies between MRIs from different centers, segmented by different core laboratories (91 % vs. 89 %).

### 3.4. Orifice annotation

As shown in Supplementary Fig. 18C, all 576 orifices and 140 valve annuli were annotated correctly (100 %) for both atria and across all modalities.

### 3.5. Division

All 140 geometries were successfully divided into the proposed regions using DIVAID. To assess potential systematic deviations of DIVAID, we first analyzed Euclidean distances between manually and automatically placed landmark points. As illustrated in Fig. 14A, discrepancies between DIVAID and experts (columns 1 and 3) were comparable to those among experts (column 2) in the LA. In fact, overall median Euclidean distances were smaller for comparisons between DIVAID and experts than for inter-expert comparisons, observed in the multi-annotator subset: 1.16 mm (0.82 – 2.87) vs. 1.82 mm (0.94 – 3.27). In the single-annotator subset, overall median Euclidean distances between DIVAID and experts were even smaller than in the multi-annotator subset, at 1.12 mm (0.80 – 2.67). Across all comparisons, landmarks *A*–*D* showed smaller median Euclidean distances than ***E***–*I*.

**Figure 14:**
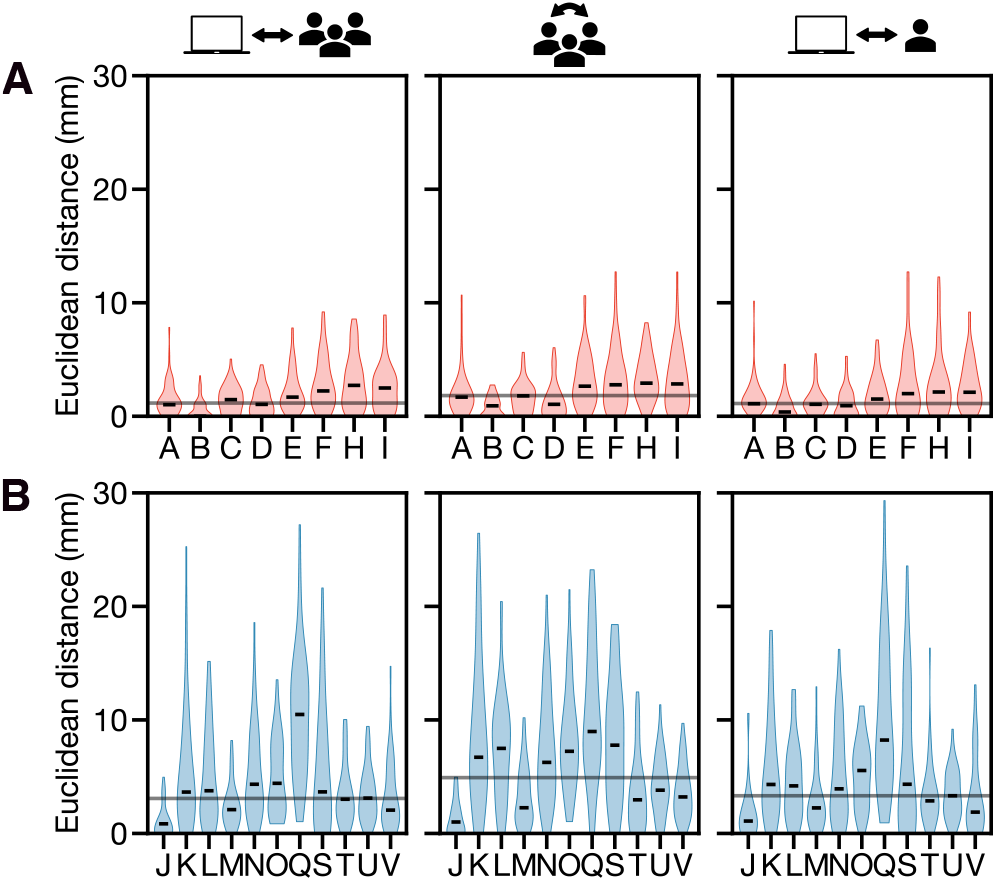
Euclidean distance between landmark points for pooled pairwise comparisons between DIVAID and experts (columns 1 and 3) as well as among experts (column 2) in the left atrium (LA) (A) and the right atrium (RA) (B). Columns 1-2 correspond to the multi-annotator subset (20 LA, 15 RA), and column 3 to the single-annotator subset (60 LA, 45 RA in total). Black lines indicate the overall median.

While discrepancies were consistently larger in the RA than in the LA, overall median Euclidean distances remained smaller for comparisons between DIVAID and experts than for inter-expert comparisons, with values of 3.08 mm (1.20 – 6.98) vs. 4.91 mm (2.12 – 8.96) in the multi-annotator subset and 3.32 mm (1.27 – 6.67) in the single-annotator subset (see Fig. 14B). Across all comparisons, the agreement was highest for landmark *J* and lowest for landmark *Q*. Analyzing individual comparisons, we found the smallest Euclidean distances for comparisons between DIVAID and expert 1, as illustrated in Supplementary Fig. 20B. Larger discrepancies were observed for landmark *K* in comparisons involving expert 2 and for landmark *S* in those involving expert 3.

Next, we investigated differences in regional surface areas obtained from either manual or automatic division, to assess if any automatically derived region was systematically smaller or larger compared to manual expert annotations. As shown in Fig. 15A, median differences in regional surface area were centered around zero across all comparisons in the LA, indicating no bias toward larger or smaller regions. We observed the smallest variability for the left pulmonary venous antrum with IQR values of –0.03 cm^2^ – 0.00 cm^2^ and –0.04 cm^2^ – 0.00 cm^2^ for comparisons between DIVAID and experts in the multi- and single-annotator subset, and 0.00 cm^2^ – 0.02 cm^2^ for inter-expert comparisons. The largest IQR values were found for the septal wall, at –0.87 cm^2^ – 0.72 cm^2^ and –0.94 cm^2^ – 1.19 cm^2^ for comparisons between DIVAID and experts in the multi- and single-annotator subset, and –0.27 cm^2^ – 3.02 cm^2^ for inter-expert comparisons. The LAA was excluded from this analysis, as its boundary had already been defined by clipping it.

**Figure 15:**
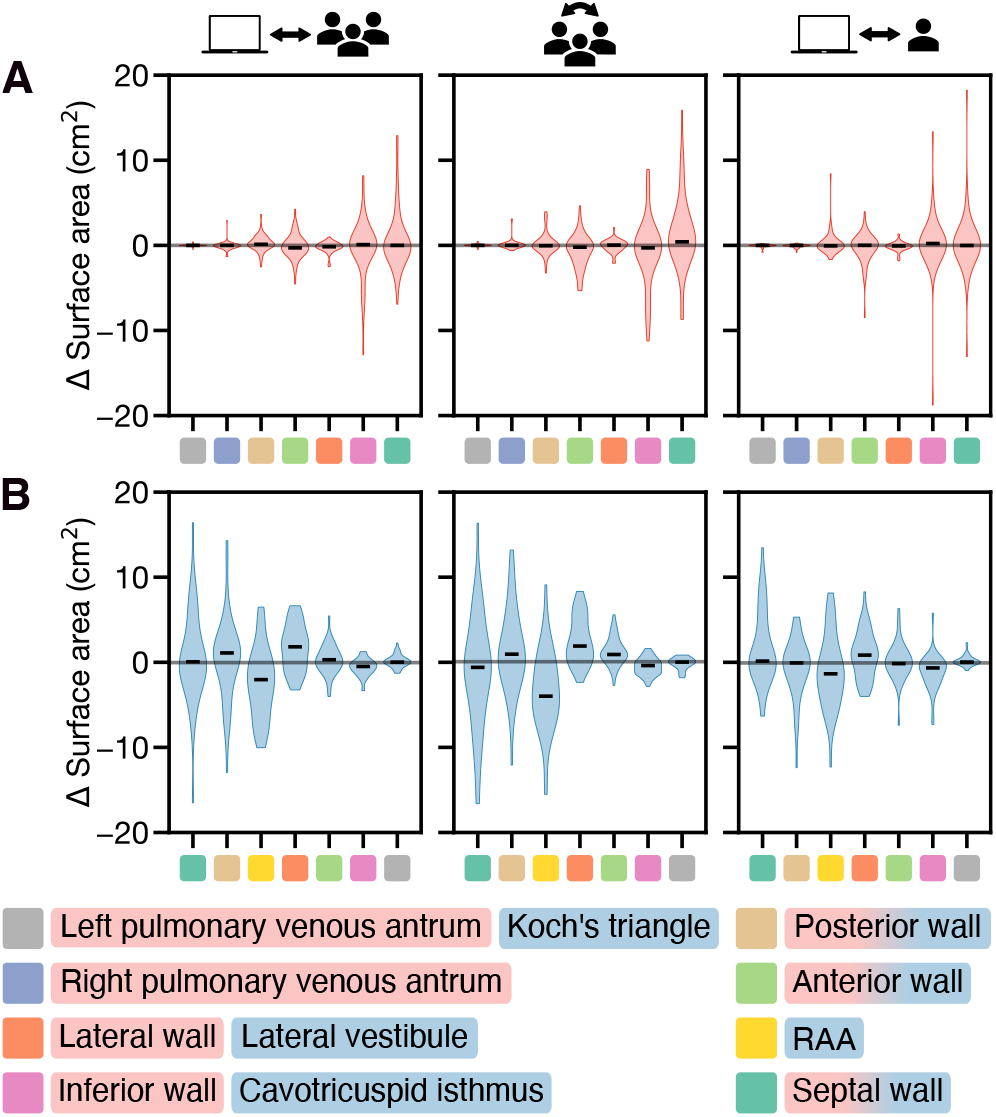
Differences in regional surface area for pooled pairwise comparisons between DIVAID and experts (columns 1 and 3) as well as among experts (column 2) in the left atrium (LA) (A) and the right atrium (RA) (B). Columns 1-2 correspond to the multi-annotaor subset (20 LA, 15 RA), and column 3 to the single-annotator subset (60 LA, 45 RA in total). Inter-expert comparisons were performed in the order 1 vs. 2, 1 vs. 3, and 2 vs. 3; this ordering determines the sign of the pooled differences. Black lines indicate the overall median. RAA, right atrial appendage.

Analogous to Euclidean distances between landmark points, differences in regional surface area were larger in the RA than in the LA (see Fig. 15B). Across all comparisons, we found the highest agreement for Koch’s triangle. The largest discrepancies were observed for the RAA and the lateral wall at –2.04 cm^2^ (–5.82 – 0.18) and –1.35 cm^2^ (–3.86 – 0.94) for comparisons between DIVAID and experts in the multi- and single-annotator subset, and –3.98 cm^2^ (–5.92 – –0.80) for inter-expert comparisons. Analyzing individual comparisons, we found that compared to DIVAID and expert 1, annotations by expert 2 and expert 3 led to larger surface areas of the RAA and the lateral wall, accompanied by smaller surface areas of the neighboring lateral vestibule, consistent with the observed trend (see Supplementary Fig. 21B). Moreover, annotations by expert 2 yielded a smaller septal wall surface area than those by the other experts, including DIVAID. The overall highest agreement was found for comparisons between DIVAID and expert 1.

We next evaluated the overlap between regions derived by DIVAID and those obtained from manual expert annotations using the DSC. As shown in Fig. 16A, median DSC values were consistently higher than 0.94 across all LA regions and comparisons. The overall median (IQR) DSC was at 0.98 (0.96 – 1.00) and 0.98 (0.96 – 1.00) for comparisons between DIVAID and experts in the multi- and single-annotator subset, and at 0.98 (0.94 – 1.00) for inter-expert comparisons. The largest overlap was observed for left the pulmonary venous antrum, while overlaps were smaller for the septal wall and the inferior wall.

**Figure 16:**
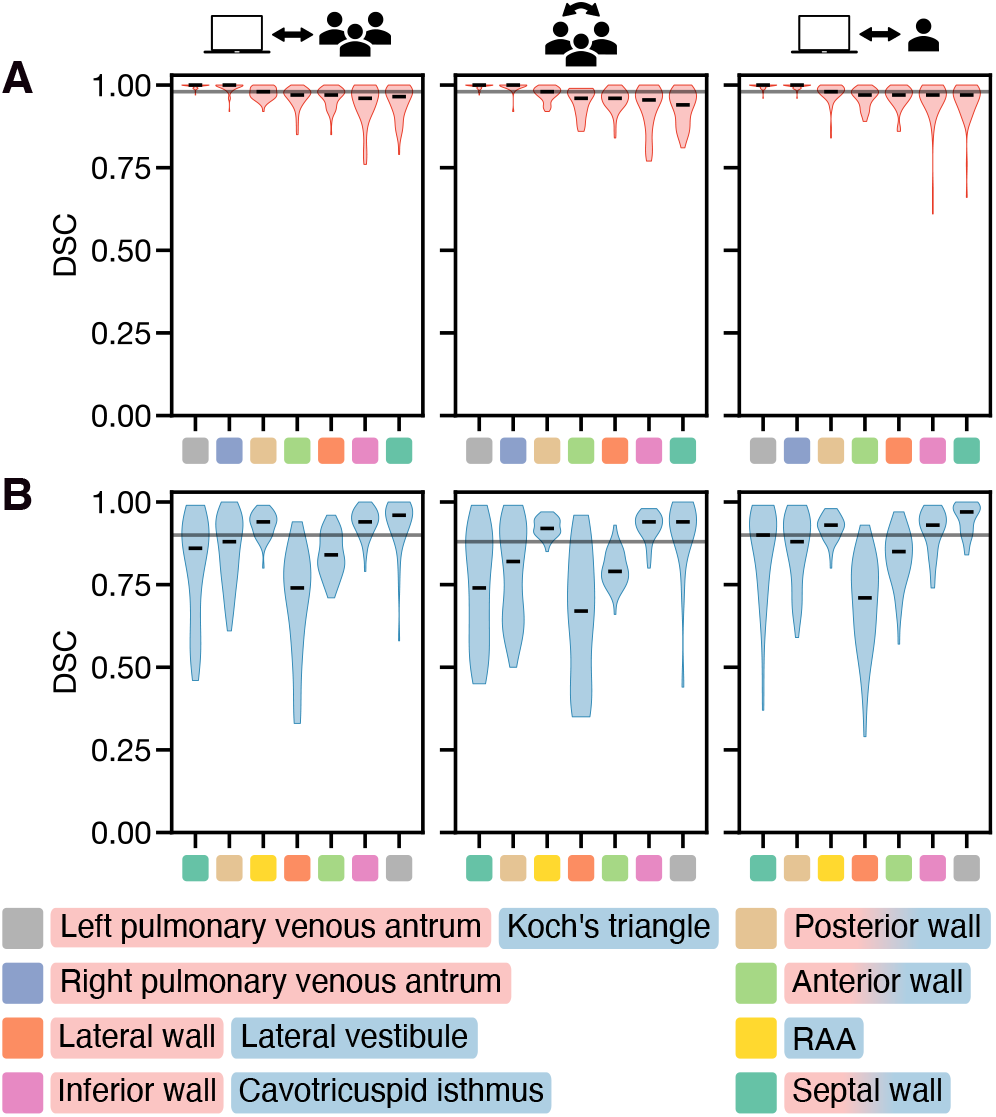
Regional Dice similarity coeffcient (DSC) for pooled pairwise comparisons between DIVAID and experts (columns 1 and 3) as well as among experts (column 2) in the left atrium (LA) (A) and the right atrium (RA) (B). Columns 1-2 correspond to the multi-annotaor subset (20 LA, 15 RA), and column 3 to the single-annotator subset (60 LA, 45 RA in total). Black lines indicate the overall median. RAA right atrial appendage.

In line with previous metrics, the overall agreement was lower in the RA compared to the LA, as illustrated in Fig. 16B. Though, the overall median (IQR) DSC was higher for comparisons between DIVAID and experts at 0.90 (0.80 – 0.95) and 0.90 (0.80 – 0.95) in the multi- and single-annotator subset than for inter-expert comparisons at 0.88 (0.74 – 0.94). The highest agreement was observed for Koch’s triangle and lowest agreement for the lateral vestibule across all comparisons. As shown in Supplementary Fig. 22B, DSC values were lower for the septal wall and the neighboring posterior venous wall in comparisons involving expert 2 than in all other comparisons. While the overall highest agreement was observed between DIVAID and expert 1 at 0.94 (0.88 – 0.98), the overall lowest agreement was found between expert 2 and expert 3 at 0.84 (0.66 – 0.92).

To assess the accuracy of regional boundaries, we computed the MASD between shortest geodesic paths derived from manually and automatically identified landmark points. As shown in Fig. 17A, the overall median (IQR) MASD was lower for comparisons between DIVAID and experts, at 0.17 mm (0.03 – 0.78) and 0.17 mm (0.03 – 0.79) in the multi- and single-annotator subset, than for inter-expert comparisons at 0.23 mm (0.05 – 0.92). Across all comparisons, the smallest median MASD was observed for paths *γ*^*^ (*A, B*) and *γ*^*^ (*C, D*), encircling the PV orifices and defining the pulmonary venous antrums. The largest MASD was observed for path *γ* ^*^ (*D, I*), with largest values for inter-expert comparisons at 1.69 mm (0.30 – 2.43).

**Figure 17:**
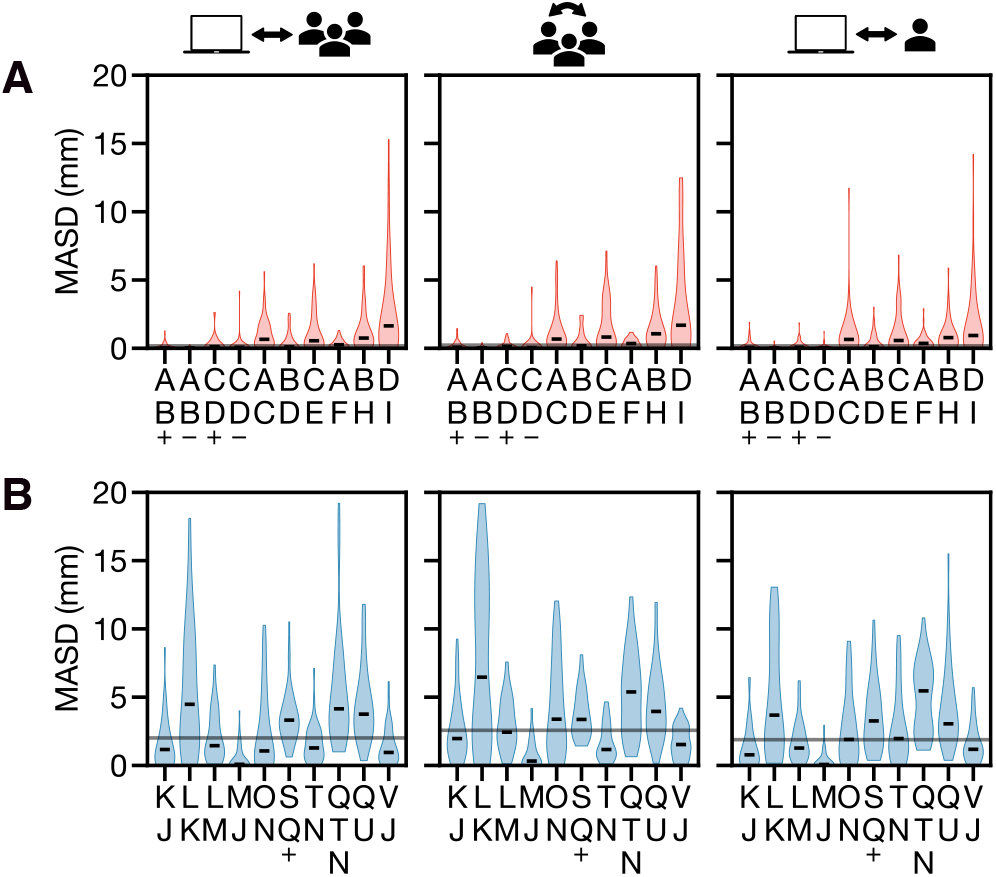
Mean average surface distance (MASD) of re-gional boundaries for pooled pairwise comparisons between DIVAID and experts (columns 1 and 3) as well as among experts (column 2) in the left atrium (LA) (A) and the right atrium (RA) (B). Columns 1-2 correspond to the multi-annotaor subset (20 LA, 15 RA), and column 3 to the single-annotator subset (60 LA, 45 RA in total). Black lines indicate the overall median. +, superior (in A) and septal (in B); -, inferior.

Consistent with previous metrics, RA geometries showed larger discrepancies than LA geometries, as shown in Fig. 17B. The overall median (IQR) MASD was 2.01 mm (0.61 – 3.87) and 1.89 mm (0.70 – 4.26) for comparisons between DIVAID and experts in the multi- and single-annotator subset, and at 2.57 mm (1.26 – 4.29) for inter-expert comparisons. Across all comparisons, path *γ* ^*^ (*M, J* ) exhibited the highest agreement. In the multi-annotator subset, the largest discrepancies were observed for path *γ*^*^ (*L, K*) at 4.48 mm (1.64 – 8.67) and 6.46 mm (1.90 – 12.76) for comparisons between DVIAID and experts and among experts. In the single annotator subset, median MASD was largest for path *γ*^*^ (*Q, T N*) at 5.46 mm (2.58 – 6.90). Analyzing individual comparisons, we found larger median MASD for paths *γ*^*^ (*L, K*) and *γ*^*^ (*O, N*) in comparisons involving expert 2 than in other comparisons, as shown in Supplementary Fig. 23. Compared to other paths, those involving landmark *Q* showed elevated median MASD across all comparisons. This is also highlighted in Supplementary Fig. 24. The overall best agreement was found for DIVAID and expert 1.

## 4. Discussion

### 4.1 Validation

In this paper, we introduced and validated the atrial division pipeline DIVAID that automatically divides bi-atrial geometries according to the recently published EHRA/EACVI consensus. DIVAID robustly preprocessed bi-atrial geometries from multiple imaging modalities and clinical centers and successfully divided them into the proposed regions, despite substantial variability in anatomy, vein configuration, surface area and volume across geometries. Our algorithm was markedly faster and more consistent than manual expert annotations, with higher Dice similarity and smaller MASD between DIVAID and experts than among experts. Released under an open-source license to ensure reproducibility, DIVAID is the first algorithm to automatically implement the newly published EHRA/EACVI consensus, enabling broad accessibility and consistent regional quantitative analyses across individuals, imaging modalities, and centers.

DIVAID provides extensive preprocessing to standardize atrial geometries and ensure consistent application of the division across diverse atrial anatomies, accounting for more than 90 % of the total computation time. By accepting input geometries with substantial anatomical variability and varying levels of preprocessing from multiple modalities, DIVAID can be applied to a wide range of atrial geometries while maintaining a high degree of automation. Existing software solutions for related tasks often rely on preprocessed input rather than incorporating such steps within their pipelines, requiring time-consuming manual operations, lacking standardization, impairing utility, and potentially aGecting the resulting analyses. Nuñez-Garcia et al. (2020) utilized a semi-automated approach to clip the veins and the LAA based on Tobon-Gomez et al. (2015a), with centerlines determined by the largest inscribed spheres. However, the need for precise seed placement at the vein tips reduces reproducibility and can lead to inconsistencies between operators.

Using our approach, more than four-fifths of all veins were automatically clipped correctly at the transition between the vein and the atrial body. Clipping success was higher for LA geometries, since RA geometries frequently exhibited short IVCs, which impeded automated clipping, as illustrated in Fig. 19. In LA geometries, the higher number of successful vein clips observed in MRI and CT datasets compared with EAMs may be attributed to the lower spatial resolution and image quality of EAMs (Nakagawa et al., 2012). The differing results for RA geometries can be explained by the frequent presence of short IVCs in CT and MRI datasets, which made precise identification of vein-to-atrial body transition challenging and limited successful clipping to approximately two-thirds of cases. Instead, EAMs typically presented longer IVC, SVC, and CS veins, which was associated with higher clipping success rates. Overall, these findings indicate that the proposed method achieves robust vein clipping across atrial chambers, imaging modalities, and centers, while offering opportunities for further methodological refinements.

The automated orifice annotation inspired by Azzolin et al. (2023); Zheng et al. (2021) demonstrated robust and accurate performance, correctly annotating all orifices across geometries regardless of varying vein and valve sizes, number of veins, or locations. Compared to existing approaches, this implementation combines high computational efficiency with flexibility, accommodating any LA geometry containing at least two PVs and one LAA. Moreover, this routine is fully automated and does not require any a-priori knowledge, such as manual labeling of specific landmarks, as in Azzolin et al. (2023); Zappon et al. (2026).

Although artificial intelligence approaches have demonstrated impressive performance in medical image segmentation (Hesamian et al., 2019), we opted for a rule-based method in DIVAID. As the 15-segment bi-atrial model provides precise definitions of characteristic landmark points and regional boundaries, we directly implemented these rules in our algorithm to ensure explainability, reproducibility and consistency across geometries and imaging modalities. Nevertheless, learning-based approaches may offer an interesting opportunity for fully automating the vein and valve clipping procedures.

Euclidean distances between landmark points and differences in surface area were comparable for DIVAID-expert and inter-expert comparisons in both, the LA and the RA. This implies that DIVAID does not introduce bias or systematic deviations from the 15-segment bi-atrial model. In fact, the agreement was higher for comparisons between DIVAID and experts than among experts across both metrics. Analyzing regional overlaps and averaged distances between boundaries, we observed similar results with higher median DSC values (LA: 0.98 vs. 0.98, RA: 0.90 vs. 0.88) and smaller median MASD (LA: 0.17 mm vs. 0.23 mm, RA: 2.01 mm vs. 2.57 mm) for DIVAID-expert than for inter-expert comparisons in the multi-annotator subset. Thus, DIVAID yields more consistent results than experts, which show larger discrepancies. Although precisely defined by Althoff et al. (2025), we observed systematic deviations of selected manual annotations from all others, as exemplified by path *γ* ^*\**^ (*L, K*) derived by expert 2. This underscores the need for further standardization and/or more elaborated definitions to achieve reliable, operator-independent quantitative regional comparisons, while highlighting the performance of DIVAID. Moreover, these results are not limited to a selected subset of geometries. Despite containing three times as many geometries (60 LA, 45 RA), the single-annotator subset showed evaluation metrics comparable to — or exceeding — those of the multi-annotator subset (20 LA, 15 RA), with median DSC of 0.98 and 0.98, and median MASD of 0.17 mm and 1.89 mm in the LA and RA, respectively. This demonstrates that DIVAID generalizes well across a wide range of diverse bi-atrial geometries from different imaging modalities and centers, achieving accuracy of regional boundaries comparable to the spatial resolution of cardiac imaging modalities.

The higher agreement observed in the LA may be attributed to its simpler, symmetric, sphere-like shape, whereas the RA exhibits a more anisotropic and complex shape with fewer prominent landmarks. Landmark points *A*–*D*, the associated paths *γ*^*\**^ (*A, B*) and *γ* ^*\**^ (*C, D*) and the related regions of the left and right pulmonary venous antrum showed the highest agreement across all comparisons. The ostia display prominent anatomical landmarks and the most superior and inferior aspects seem to be easily consistently identifiable, both for the experts and for DIVAID. Similar results were found for Koch’s triangle in the RA, defined by the most inferior aspect of the CS and the clock model of the TV.

In contrast, the largest discrepancies observed across all metrics were associated with landmark *Q* in the RA. Along with landmark *P*, these are the only landmarks not located on an orifice. The lack of prominent anatomical landmarks in this area, combined with the inherent variability of the RAA, renders consistent identification of these landmarks particularly challenging, highlighting the need for consistent and automated annotation approaches. In this context, the importance of analyzing overlap-based metrics in combination with boundary-based metrics becomes evident (Maier-Hein et al., 2024). The RAA and the lateral wall are separated from the lateral vestibule by the shortest geodesic path *γ*^*^ (*Q, T N*), which showed a high discrepancy across annotations. However, while this finding also resulted in a lower DSC for the lateral vestibule, the median DSC of the RAA and the lateral wall remains highest among RA regions. This can be explained by the substantial difference in size between both regions, affecting DSC values despite normalization.

### 4.2 Limitations

Despite the presented accordance with manual expert annotations, DIVAID can still be further improved. Though challenging to identify automatically, particularly in EAMs, automated clipping of the valves would be advantageous. Moreover, automated placement of seed nodes on salient sub-structures would increase both the efficiency and the level of automation of DIVAID. However, as illustrated above, this may remain complex for short veins or veins not fully represented in the geometries. The algorithm expects LA geometries to exhibit an MV annulus, an LAA and at least two PVs, and RA geometries to exhibit a TV annulus, an SVC, an IVC and a CS. For geometries lacking these features, orifices need to be estimated manually. Dijksta’s algorithm produces a one-dimensional path along mesh edges, which enables separation of surface-mesh components but is not directly applicable to volumetric meshes. However, DIVAID could be enhanced by projecting shortest geodesic paths along edge normals derived from adjacent elements. Lastly, we aim to extend DIVAID by atrial coordinates aligned with regional boundaries, thereby transforming the discretized 15-segment bi-atrial model into a fully parametrized framework. This extension would enable data mapping across geometries, as well as 2D data visualization, as shown by Roney et al. (2019). Moreover, it would further enhance standardized data acquisition and precise clinical analyses, and support their integration into cardiac digital twin generation pipelines (e.g., fibrosis distribution, fiber architecture or region-specific cellular properties).

## 5. Conclusion

Here, we present DIVAID, the first software solution to automatically divide bi-atrial geometries according to the recently published EHRA/EACVI consensus. Released under an open-source license for reproducibility and adoption, DIVAID demonstrates robust preprocessing of geometries despite substantial anatomical variability and shows high agreement with manual annotations, while outperforming experts in both speed and consistency. By enabling automated and consistent atrial regionalization without requiring expert knowledge, DIVAID lays the foundation for large-scale standardized analyses of regional atrial data across patients, imaging modalities, studies, centers, and computational models. The integration of such harmonized, multi-dimensional datasets through data-driven approaches holds the potential to unlock key insights for advancing atrial arrhythmia research and personalized treatment strategies.

## Data Availability

All data produced in the present study are available upon reasonable request to the authors. The code is available online at

https://gitlab.kit.edu/kit/ibt-public/divaid

## Declaration of generative AI and AI-assisted technologies in the manuscript preparation process

During the preparation of this work, the authors used ChatGPT in order to rephrase individual sentences for clarity. After using this tool/service, the authors reviewed and edited the content as needed and take full responsibility for the content of the published article.

## CRediT authorship contribution statement

**Christian Goetz:** Conceptualization, Data curation, Investigation, Methodology, Software, Validation, Visualization, Writing – original draft. **Martin Eichenlaub:** Data curation, Validation. **Kerstin Schmidt:** Validation. **Felix Wiedmann:** Funding acquisition, Validation. **Eric Invers Rubio:** Data curation, Validation. **Patricia Martínez Díaz:** Conceptualization, Methodology, Software. **Armin Luik:** Validation. **Till Althoff:** Data curation, Validation. **Constanze Schmidt:** Funding acquisition, Supervision. **Axel Loewe:** Conceptualization, Funding acquisition, Methodology, Software, Supervision, Writing – original draft.

## 6. Supplementary material

### 6.1 Validation of preprocessing

**Figure 18:**
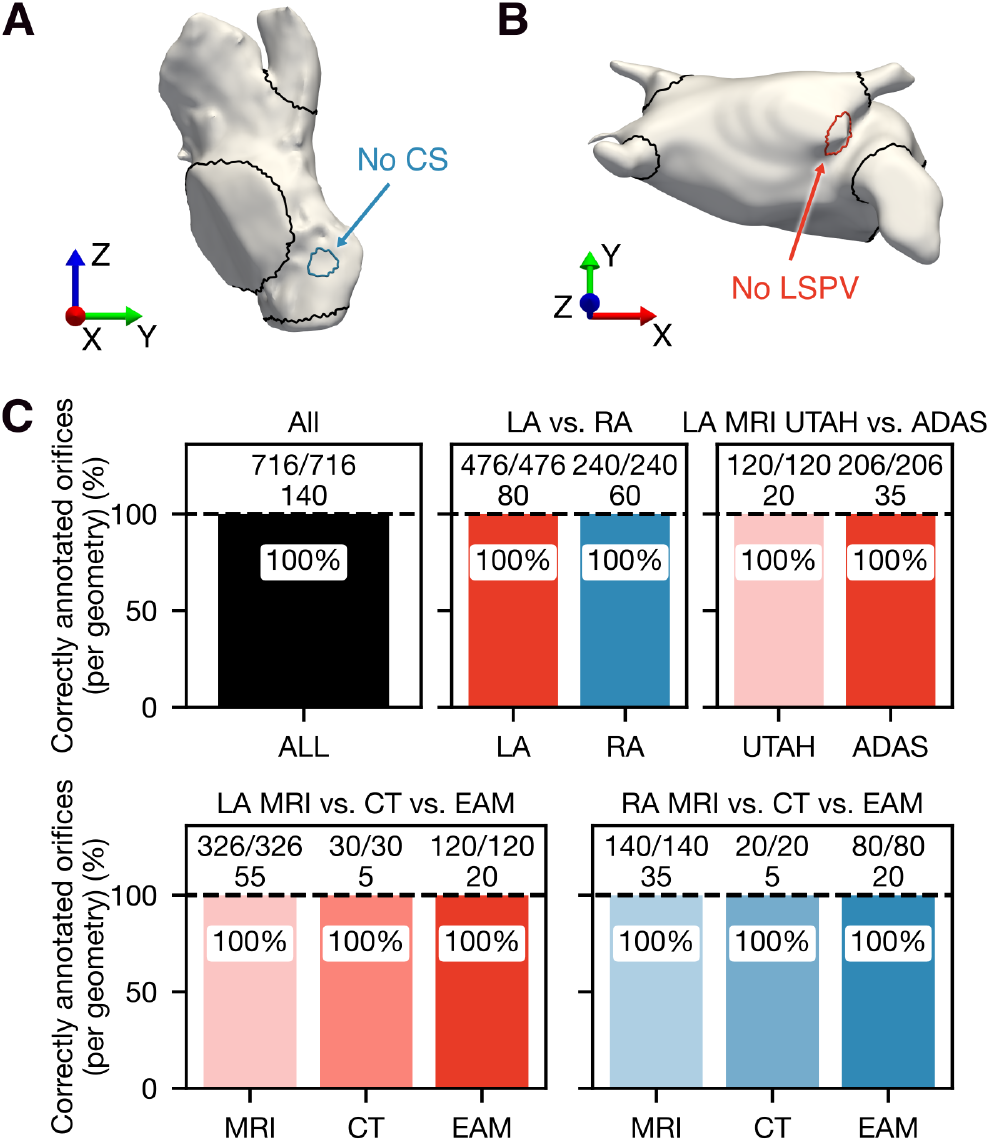
Validation of the preprocessing. (A) Right atrial (RA) geometry without the coronary sinus (CS) incorporated. (B) Left atrial (LA) geometry without the left superior pulmonary vein (LSPV) incorporated. (C) Correctly annotated orifices per geometry. In each panel, the upper row corresponds to the number of correctly annotated orifices and the lower row to the number of geometries. MRI, magnetic resonance imaging; CT, computed tomography; EAM, electroanatomical mapping.

**Figure 19:**
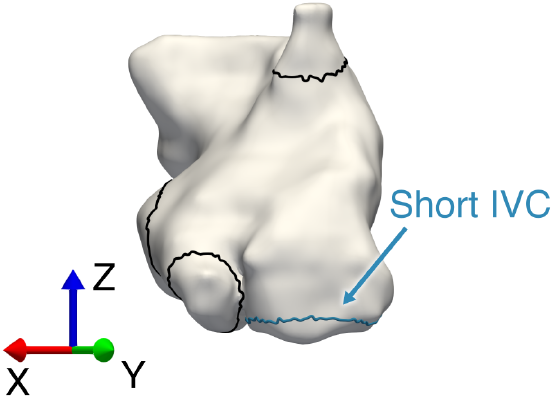
Representative geometry with a short inferior vena cava (IVC), resulting in increased difficulty for automated vein clipping.

### 6.2 Validation of division – individual comparisons

**Figure 20:**
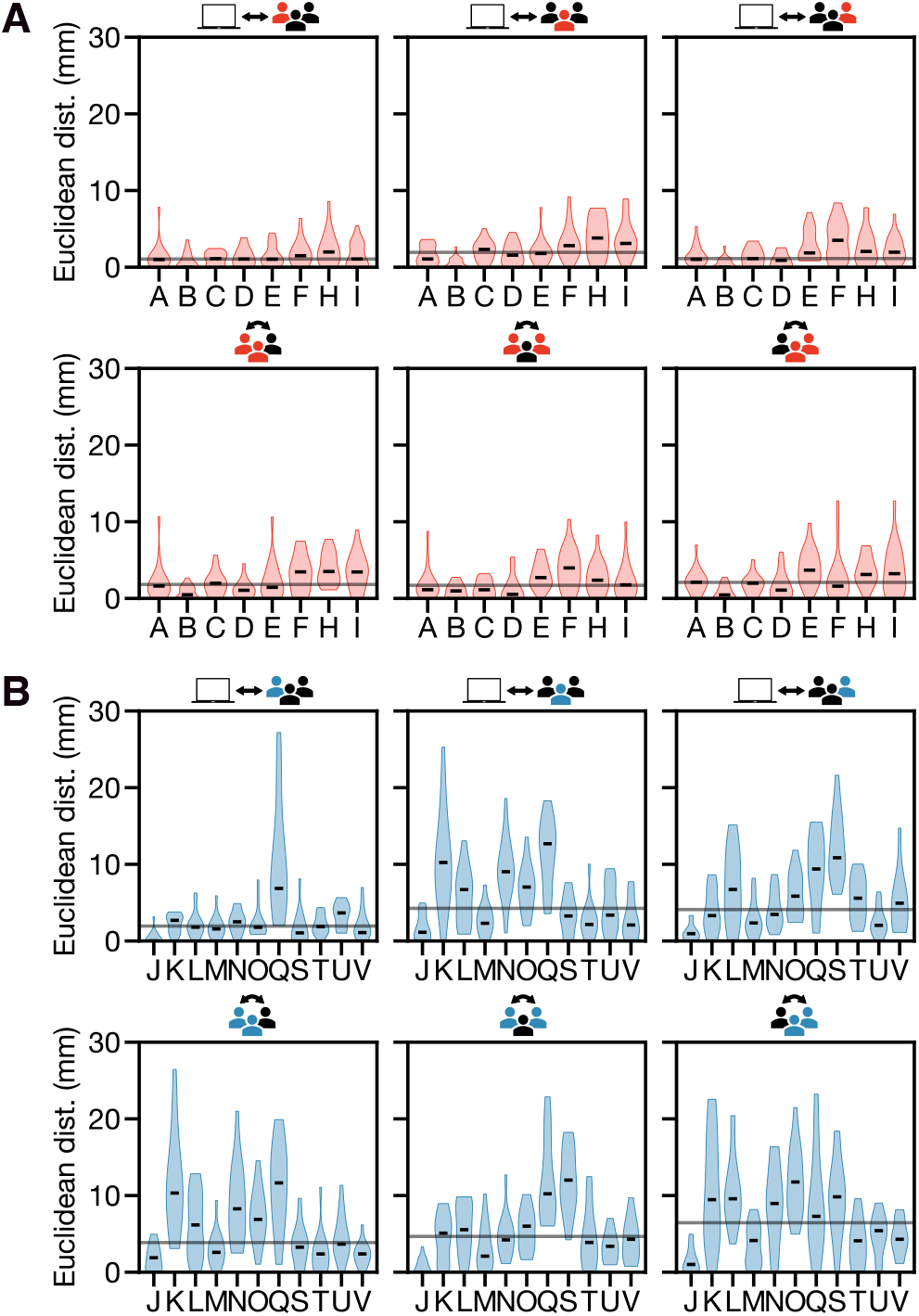
Euclidean distance between landmark points for pairwise comparisons between DIVAID and experts (first row) as well as among experts (second row) in the left atrium (LA) (A) and the right atrium (RA) (B) using the multi-annotaor subset (20 LA, 15 RA). Black lines indicate the overall median. Dist., distance.

**Figure 21:**
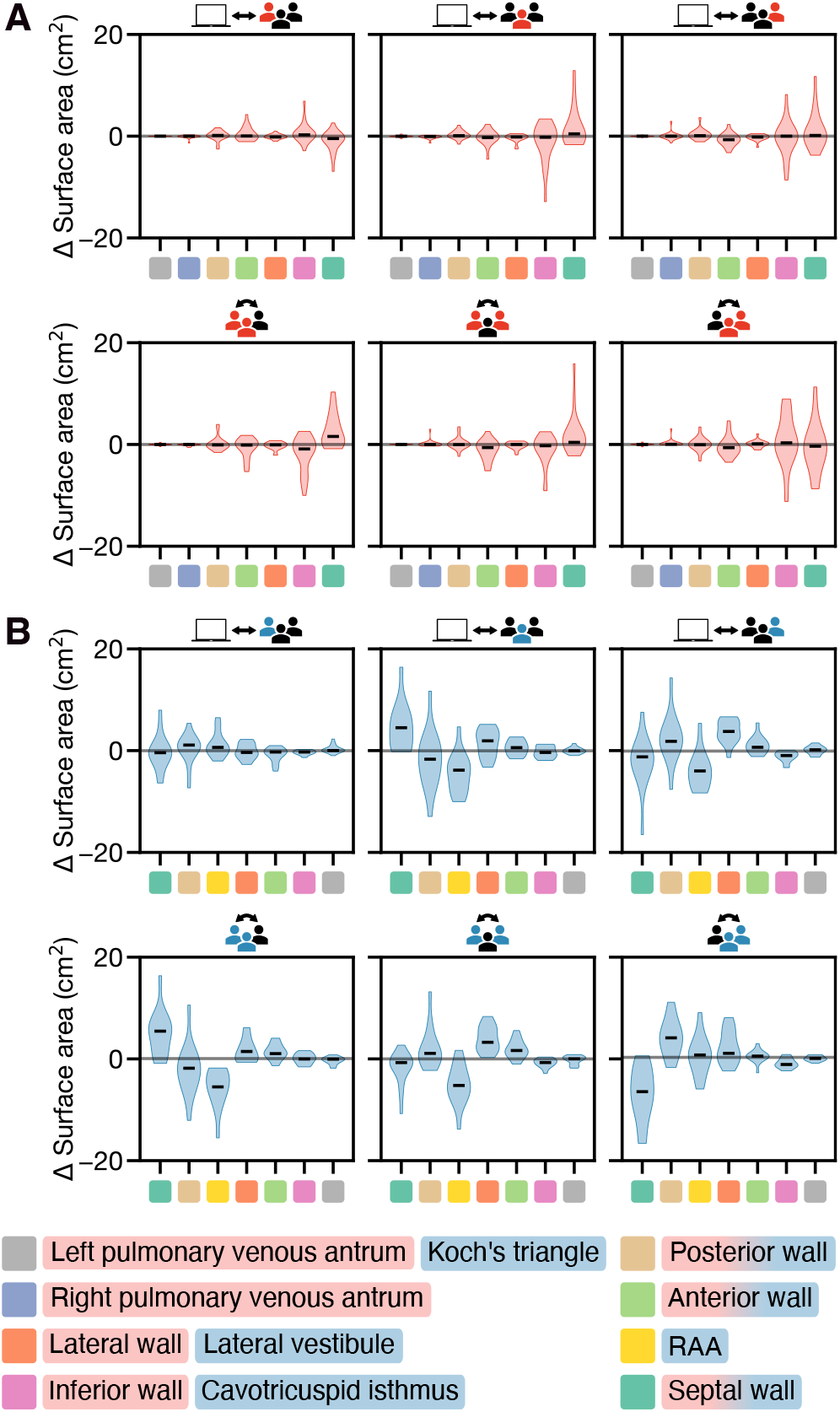
Differences in regional surface area for pair-wise comparisons between DIVAID and experts (first row) as well as among experts (second row) in the left atrium (LA) (A) and the right atrium (RA) (B) using the multi-annotaor subset (20 LA, 15 RA).

**Figure 22:**
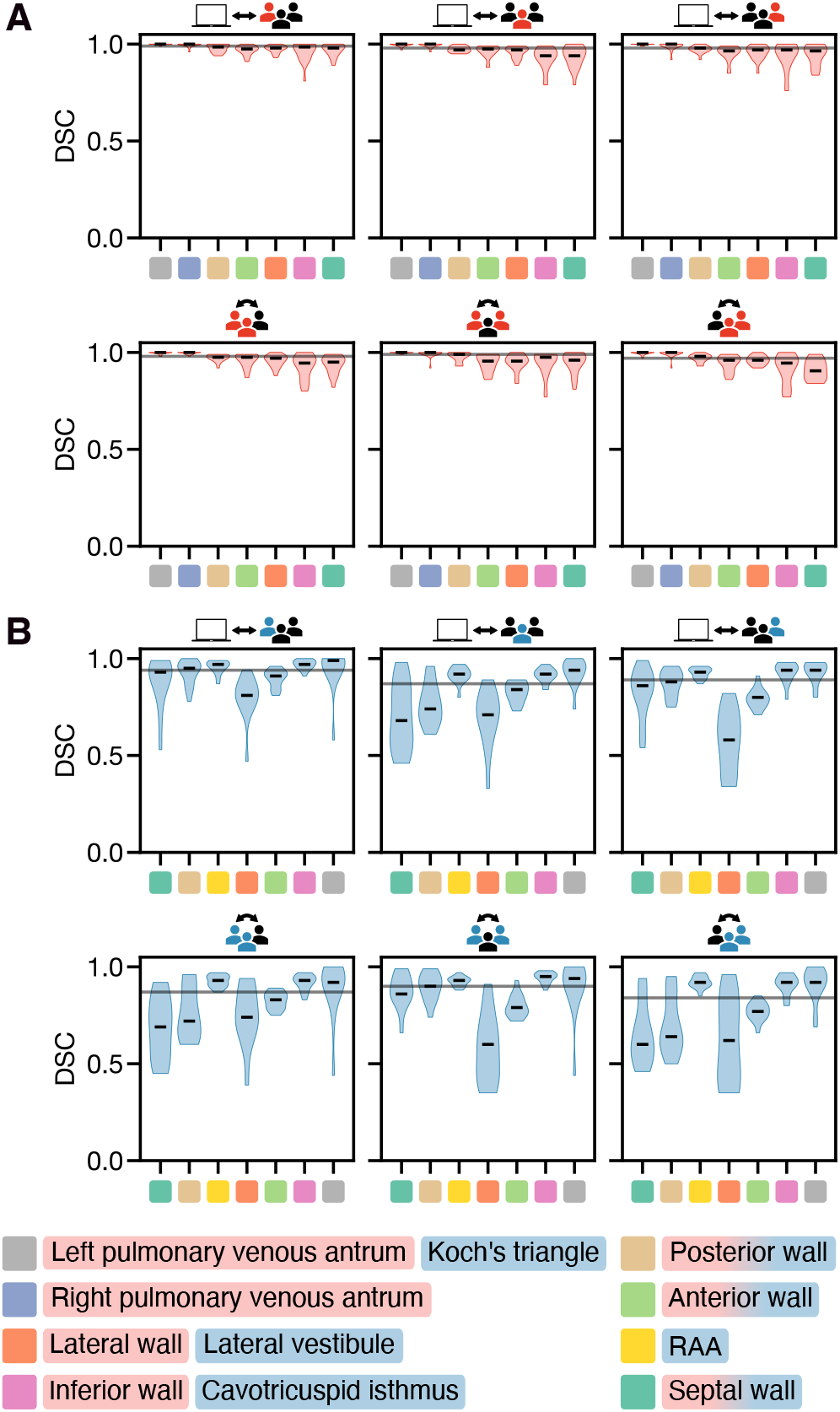
Regional Dice similarity coeAcient (DSC) for pairwise comparisons between DIVAID and experts (first row) as well as among experts (second row) in the left atrium (LA) (A) and the right atrium (RA) (B) using the multi-annotaor subset (20 LA, 15 RA). Black lines indicate the overall median. RAA, right atrial appendage.

**Figure 23:**
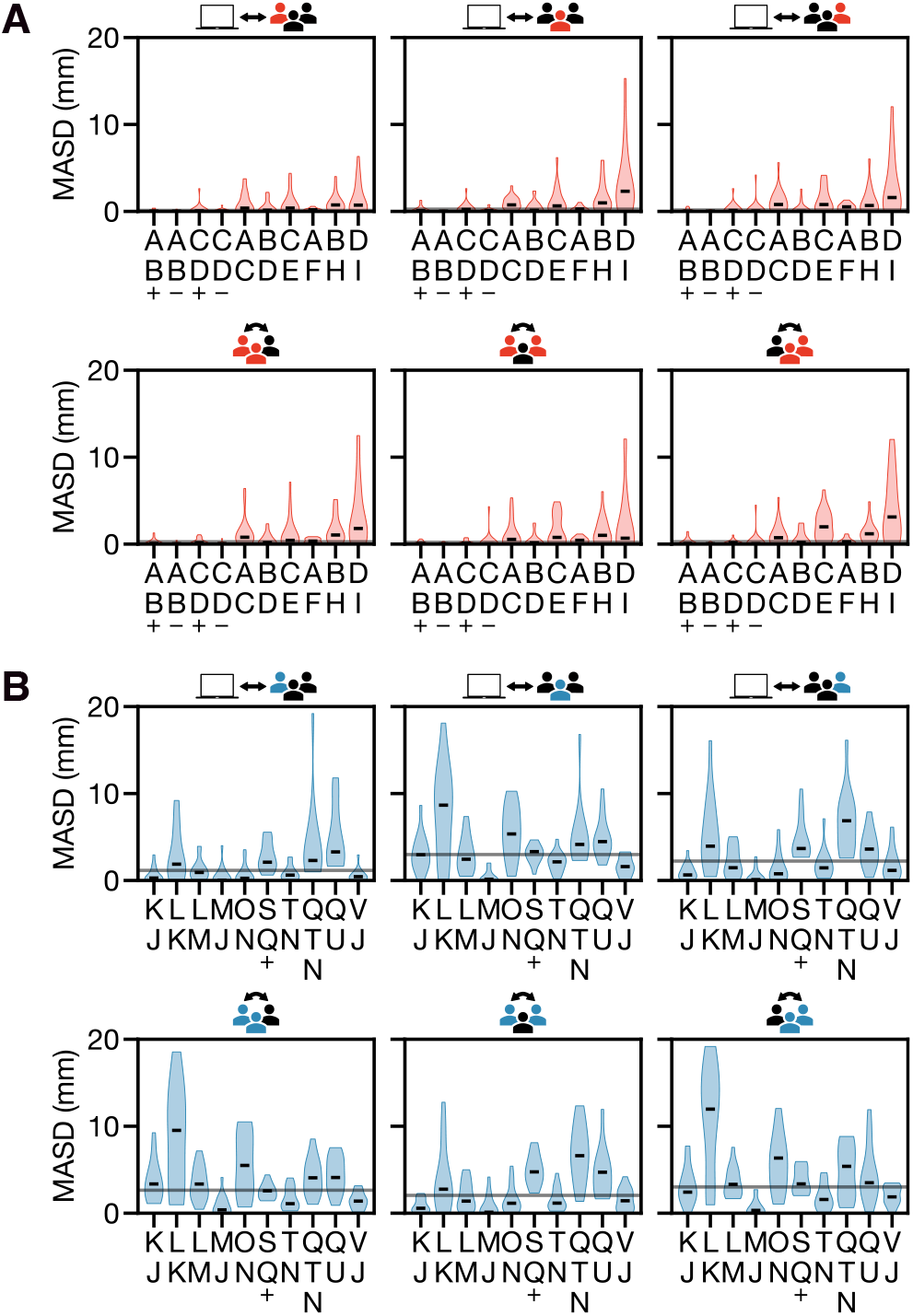
Mean average surface distance (MASD) of regional boundaries for pairwise comparisons between DIVAID and experts (first row) as well as among experts (second row) in the left atrium (LA) (A) and the right atrium (RA) (B) using the multi-annotaor subset (20 LA, 15 RA). Black lines indicate the overall median. +, superior (in A) and septal (in B); -,inferior.

**Figure 24:**
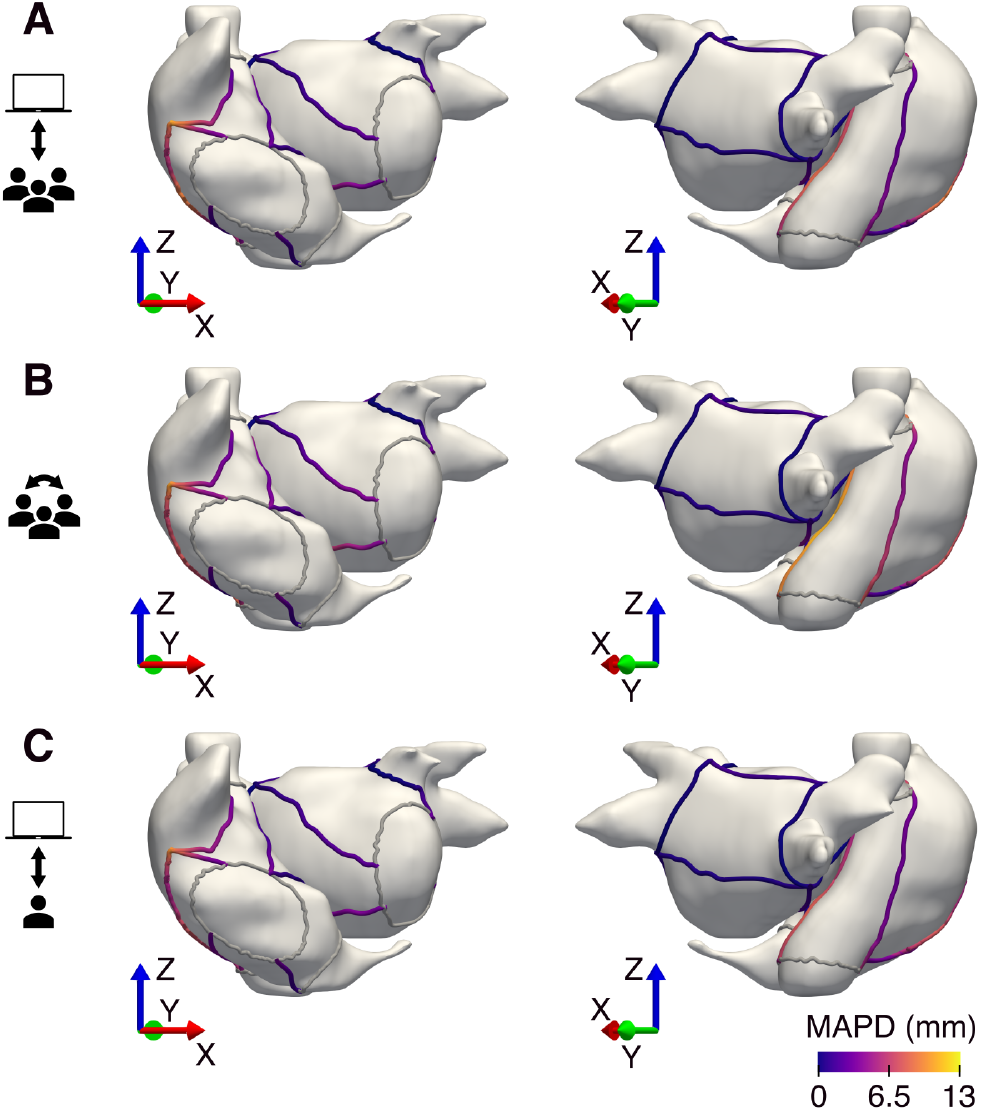
Mean average parametrized distance (MAPD) of regional boundaries for pooled pairwise comparisons between DIVAID and experts (first and third row) as well as among experts (second row). Analogous to the MASD, for each node 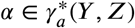, we compute the shortest Euclideen distance to any node in 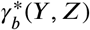. Linear interpolation between nodes defines a continuous, parametrized distance function along 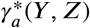. The same procedure is applied to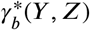 and the mean of both directions is computed. Averaging across all geometries within a given subset yields the MAPD.

### 6.3. Validation metrics

#### Metrics Reloaded toolkit report

This report summarizes the outputs of the Metrics Reloaded online toolkit hosted at https://metrics-reloaded.dkfz.de. It highlights the questions asked and their descriptions, as well as user responses and recommended metrics.

We kindly ask you to refer to the main article of this toolkit when using the results for publishing. You can visit https://metrics-reloaded.dkfz.de/publications for more information.

#### Problem category

***Semantic Segmentation***

Suggested metrics

1. Overlap-based Metric: <u>Dice Similarity Coefficient (DSC)</u>
2. Boundary-based Metric: <u>Mean Average Surface Distance (MASD)</u>

Steps:

1. **Description:** Welcome to metric selection for Semantic Segmentation Throughout this toolkit you will be asked a number of questions, from different processes. Based on your answers, the toolkit will determine the most relevant metrics for your research problem.
2. **Description:** Selecting overlap-based segmentation metrics (if any; S6) In segmentation problems, counting metrics such as the DSC or IoU measure the overlap between the reference annotation and the prediction of the algorithm. As they can be considered the de facto standard for assessing segmentation quality and are well-interpretable, we recommend using them by default unless the target structures are consistently small (relative to the grid size) and the reference may be noisy. Repeat for each class: If problem fingerprints differ between classes (e.g., simultaneous segmentation of convex and tubular structures), a class-specific metric pool must be generated (background class: optional).
3. **Question:** Does your data feature consistently small structures relative to pixel size? **Answer:** No **Description:** Small size of structures relative to pixel size

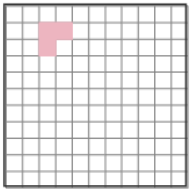 Structures of the provided class are only a few pixels in size. *Example: Multiple sclerosis lesions in magnetic resonance imaging (MRI) scans*. *Note: For further information please refer to FP3.1 in our article: Maier-Hein et al. 2024.*
4. **Question:** Is there exclusive interest in structure centers in your research? **Answer:** No **Description:** Particular importance of structure center (e.g. in cells, vessels)

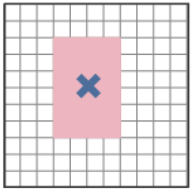 The biomedical application requires accurate knowledge of structure centers. *Example: Cell centers are subsequently used for cell tracking and cell motion characterization, so false center movement should be suppressed*. *Note: For further information please refer to FP2.3 in our article: Maier-Hein et al. 2024*.
5. **Question:** According to the this information, do class confusions, i.e., over-vs. undersegmentation, have equal or unequal weight in your study? **Answer:** Equal handling **Description:** Penalization of errors: Unequal severity of class confusions

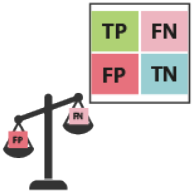 Any class can be confused with another, but certain mismatches are more severe than others, from a domain point of view. This holds especially true (1) in screening tasks, in which FN are typically more severe than FP, (2) in retrieval tasks, in which FP are typically more severe than FN and (3) in tasks with ordinal rating. Note that class confusions in this context can be considered as costs for individual cells in the confusion matrix, while “interest across classes” (FP2.5.1) would consider all matrix cells related to one class as a whole. It is important to note that this fingerprint only considers “a priori costs” of a task that are *irrespective of the class prevalences in the data*. This distinction is necessary, because one can also tweak the confusion costs in hindsight to compensate for certain imbalances in the data (not considered here). Class confusions in this context can be considered as costs for individual cells in the confusion matrix, while “interest across classes” would consider all matrix cells related to one class as a whole. *Example 1 (binary, OD): Polyp detection; a FN (missed polyp) is clinically much more severe than a FP*. *Example 2 (multi-class): Depending on the application, confusing different kinds of immune cells is more problematic compared to confusing an immune cell with a tumor epithelial cell*. *Example 3 (multi-class): Lung tumor categorization T1-T5 depends largely on structure size, implying an ordinal scale of classes. Thus, penalization of class confusions should reflect this ordinal scale*. *Note: For further information please refer to FP2.5.2 in our article Maier-Hein et al. 2024*.
6. **Question:** According to this information, which metric better suits your problem? **Answer:** Dice Similarity Coefficient (DSC) **Description:** Dice Similarity Coefficient (DSC) vs. Intersection over Union (IoU) <u>Dice Similarity Coefficient (DSC):</u>
  ∼ Identical to F1 Score
  ∼ Close relation to IoU
  ∼ Preference in medical community <u>Intersection over Union (IoU):</u>
  <u>Identical to Jaccard Index</u>
  ∼ Close relation to DSC
  ∼ Preference in computer vision community
  *Note: For further information please refer to DG6.1 in our article: Maier-Hein et al. 2024*
7. **Description:** Selecting an overlap-based segmentation metric completed As overlap-based segmentation metric, Dice Similarity Coefficient (DSC) has been added to the metric candidates pool. You are ready for the next step! If problem fingerprints differ between classes (e.g., simultaneous segmentation of convex and tubular structures), a class-specific metric pool must be generated (background class: optional). Add the selected metric(s) to class-specific metric pool.
8. **Description:** Selecting boundary-based segmentation metrics (if any; S7) Key weaknesses of overlap-based metrics include shape unawareness and limitations when dealing with small structures or high size variability. Our general recommendation is therefore to complement an overlap-based metric with a boundary-based metric. Repeat for each class: If problem fingerprints differ between classes (e.g., simultaneous segmentation of convex and tubular structures), a class-specific metric pool must be generated (background class: optional).
9. **Question:** Are there overlapping or touching target structures in your data? **Answer:** No **Description:** Possibility of overlapping or touching target structures (e.g. medical instruments or cells)

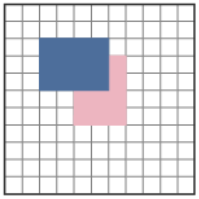 Different instances of a class can overlap or touch each other. *Examples: Overlapping cells or organisms, such as BBBC010 (worms in a dish); overlapping medical instruments in laparoscopy*. *Note: For further information please refer to FP3.5 in our article Maier-Hein et al. 2024*.
10. **Question:** Is there particular importance of structure boundaries in your research? **Answer:** Yes **Description:** Particular importance of structure boundaries

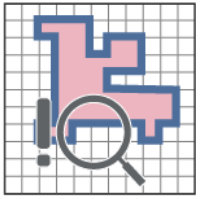 The biomedical application requires exact structure boundaries. *Example: Segmentation for radiotherapy planning; knowledge of exact structure boundaries is crucial to destroy the tumor while sparing healthy tissue*. *Important: Overlap-based metrics do not measure shape agreement. In the cases of complex shapes (high boundary-to-volume ratio) it is therefore typically advisable to set this property to TRUE*. *Note: For further information please refer to FP2.1 in our article Maier-Hein et al. 2024*.
11. **Question:** Is compensation for annotation imprecisions requested in your research? **Answer:** No **Description:** Penalization of errors: Compensation for annotation imprecisions requested The reference annotation is typically only an approximation of the (forever unknown) ground truth. It may be desirable to compensate for known uncertainties, such as intra-rater or inter-rater variability, by configuring the metric accordingly.

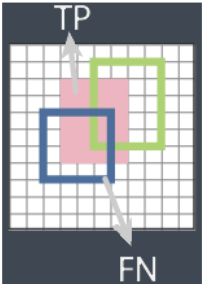 This is only possible for some metrics. *Note: For further information please refer to FP2.5.7 in our article Maier-Hein et al. 2024*.
12. **Question:** Are there spatial outliers in the reference annotation in your data? **Answer:** No **Description:** Uncertainties in the reference: Possibility of spatial outliers in reference annotation

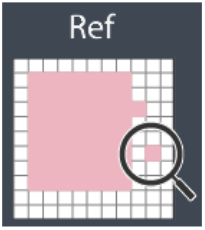 The reference may feature spatial outliers that are distant from the (unknown) ground truth. *Note: For further information please refer to FP4.3.2 in our article Maier-Hein et al. 2024*.
13. **Question:** Which type of handling of spatial outliers fits your problem more? **Answer:** Distance-based penalization with whole contour focus **Description:** Penalization of errors: Handling of spatial outliers

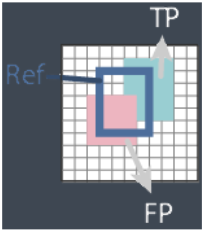 Spatial outliers are FP predictions that feature a large distance to the reference. They can be handled in three different ways: **Distance-based penalization with outlier focus:** Individual outliers should be heavily penalized as a function of the distance to the reference contour. **Distance-based penalization with whole contour focus:** Outliers should be penalized as a function of the distance to the reference, but the assessment should focus on the general contour agreement rather than individual outliers. **Existence-based penalization:** The existence of spatial outliers should be penalized irrespective of their distance to the reference contour. Note that distance-based penalization is not possible when either the reference or the prediction is empty. In applications in which many of such cases potentially occur, we therefore recommend an existence-based penalization. *Note: For further information please refer to FP2.5.6 in our article Maier-Hein et al. 2024*.
14. **Question:** According to this information, which metric better suits your problem? **Answer:** Mean Average Surface Distance (MASD) **Description:** Mean Average Surface Distance (MASD) vs. Average Symmetric Surface Distance (ASSD) <u>Mean Average Surface Distance (MASD):</u>
  ∼ Equal contribution of reference and prediction boundaries to the metric score
  - Possibility of misleading results in corner cases (e.g. tiny prediction closely located to the reference)
  <u>Average Symmetric Surface Distance (ASSD):</u>
  - Domination of the metric score by the larger contour
  *Note: For further information please refer to DG7.2 in our article: Maier-Hein et al. 2024*
15. **Description:** Selecting a boundary-based segmentation metric completed As boundary-based segmentation metric, Mean Average Surface Distance (MASD) has been added to the metric candidates pool. If problem fingerprints differ between classes (e.g., simultaneous segmentation of convex and tubular structures), a class-specific metric pool must be generated (background class: optional). Add the selected metric(s) to class-specific metric pool. You are ready for the next step!
16. **Description:** Additional metrics for your study In addition to the recommended metrics, we encourage you to select further additional metrics specific to your problem if needed<u>.Application-specific metrics:</u>The pool of standard metrics can be complemented with custom metrics to address application-specific complementary properties. For example, in liver segmentation, the absolute volume of the segmented liver is an important clinical parameter, so the Absolute Volume Error could be added to the metric pool<u>.Non-reference-based metrics</u>:Validation and evaluation of algorithms could go far beyond purely technical performance. Thus, non-reference-based metrics assessing speed, memory consumption or carbon footprint, for example, can be added to the metric pool.
17. **Description:** The Metric Selection toolkit completed The metric pool has been generated. It can then be complemented by application-specific metrics (e.g. absolute volume difference if the exact volume is of particular interest) as well as non-reference-based metrics (assessing run time or carbon footprint, for example). In the final stage, it is necessary to apply the selected metrics to the designated dataset. You can check the <u>Metric Application</u> page for additional guidance. You can use this report as attachment to your publication. Created on: Thu Jul 24 2025 15:56:20 GMT+0200 (Central European Summer Time)

